# Combined Metabolic and Microstructural Tractometry of the Superior Longitudinal Fasciculus in Healthy Brains: A Proof-of-Concept Study

**DOI:** 10.64898/2026.08.25.26361054

**Authors:** Archith Rajan, Sourav Bhaduri, Subhanon Bera, Laiz Laura de Godoy, Mauro Hanaoka, Sulaiman Sheriff, Madhura Ingalhalikar, Laurie A. Loevner, Suyash Mohan, Sanjeev Chawla

**Author notes:** Address correspondence to: Sanjeev Chawla, PhD, DABMP Research Associate Professor, Perelman School of Medicine at the University of Pennsylvania, Philadelphia, PA-19104, United States; Suyash Mohan, MD, PDCC, Professor of Radiology & Neurosurgery, Perelman School of Medicine at the University of Pennsylvania, Philadelphia, PA-19104, United States.

## Abstract

**Introduction:** The superior longitudinal fasciculus (SLF) is a major association fiber bundle implicated in cognition, visuospatial attention, language, and motor control, and its impairment is linked to several neurological and neuropsychiatric disorders. This proof-of-concept study was performed with three main objectives in healthy adults. First, to fuse whole brain spectroscopic (WBSI) and diffusion MRI (dMRI) derived parametric maps along the SLF I and II segments to quantify their spatial concordance, second, to evaluate regional metabolite concentrations and microstructural properties along these trajectories and finally, to determine the relationships between the WBSI and dMRI parameters within these segments.

**Methods:** Ten healthy adults (4F, 6M; mean age 31.4 ± 7.53 years) underwent 3T MRI including multi-shell high angular resolution diffusion imaging and WBSI. After preprocessing and non-linear co-registration, WBSI-derived white matter metabolite maps and neurite orientation dispersion and density imaging (NODDI) / diffusion tensor imaging (DTI) derived parametric maps were spatially aligned and projected along the centroid of reconstructed SLF I and II segments divided into 20 discrete, anatomically contiguous sections.

**Results:** A strong spatial alignment between WBSI and dMRI imaging modalities was confirmed by mutual information and Pearson’s correlation analyses. Intra-subject repeatability, as assessed from a single participant scanned three times, demonstrated high tract reconstruction reliability (mean Dice similarity coefficients >0.79; track density-weighted Dice >0.97) and acceptable intra-subject coefficients of variation. Inter-subject coefficients of variation were within acceptable ranges (∼3-17%) for most parameters, with free water fraction (fiso) exhibiting relatively higher variability. Single and multivariate regression analyses revealed significant associations between WBSI and dMRI tract profiles: choline/creatine (Cho/Cr) and choline/ N-acetyl aspartate (Cho/NAA) ratios showed positive linear associations with intra-cellular volume fraction (ficvf) and fractional anisotropy (FA), and negative associations with mean diffusivity (MD) along bilateral SLF I, with ficvf and MD identified as the strongest combined predictors of metabolite ratios.

**Conclusion:** Co-localization/fusion of WBSI and NODDI/DTI data into one framework offers a reliable, user-independent way for mapping regional metabolite and microstructural alterations along the path of SLF. Moving forward, this image processing pipeline has the potential to enhance diagnosis and clinical assessment of neurological disorders linked to SLF damage.

## Introduction

Superior longitudinal fasciculus (SLF) is the largest associative fiber bundle connecting frontal and parietal brain regions. The SLF is involved in several vital normal functions such as cognition, visuospatial attention and memory, motor control, and comprehension of language (Wang et al., 2016). Impairment of the SLF is associated with several neurological, neurodevelopmental, and neuropsychiatric disorders including schizophrenia (Karlsgodt et al., 2008), traumatic brain injury (Bendlin et al., 2008), Alzheimer’s disease (Tahara-Eckl et al., 2023), Autism(Fitzgerald et al., 2018),and Parkinson’s disease (Chen et al., 2018). Therefore, the development of non-invasive biomarkers for assessing neurochemical and microstructural profiles of SLF is vital to better understand the underlying pathophysiology of these conditions and potentially develop the targeted therapies.

Using single voxel or multivoxel proton MR spectroscopy (1H-MRS) and diffusion MRI (dMRI) techniques independently, several previous studies have reported metabolic and microstructural alterations from multiple brain regions of patients with neurological disorders in which SLF is known to be compromised (Catani et al., 2003; Cho et al., 2008; Michelutti et al., 2025; Su et al., 2016). However, combining 1H-MRS and dMRI within an integrated analytical framework offers a more comprehensive and reliable approach to understanding pathogenesis and progression of these neurological disorders (Irwan, Sijens, Potze, & Oudkerk, 2005; A. A. Maudsley et al., 2015; Zecca, Palombelli, Vanacore, & Canese, 2026). However, a key limitation of previous combination studies was that the regions of interest were not spatially concordant between 1H-MRS and diffusion MRI modalities, which prevented the synergistic integration of parameters from similar regions (Manning et al., 2017). This spatial misregistration can be attributed to inherent limitations of conventional single-voxel or multivoxel spectroscopy sequences, which are constrained by restricted spatial coverage and poor spatial resolution.(Tomiyasu & Harada, 2022; Weingartner et al., 2022) These technical shortcomings prevent an exact anatomical match between the 1H-MRS and other imaging modalities. Additionally, the manual selection of anatomical regions typically relies on subjective and potentially inconsistent judgments about the anatomical landmarks, increasing the probability of error in obtaining data from the same anatomical region in cross-sectional and longitudinal studies. This approach precludes simultaneous analysis of metabolite profile and microstructural integrity from similar brain regions under normal and pathological conditions. For instance, significant differences in diffusion tensor imaging (DTI) derived parameters were observed between normal controls and schizophrenia patients from brain regions along the path of SLF in a study, but no corresponding changes in metabolite pattern between the two groups were found (Rowland et al., 2009). These findings may suggest dissociation between structural and metabolic brain abnormalities in schizophrenia and as such changes in white matter integrity as revealed by DTI may not always be accompanied by detectable changes in metabolite concentrations. Alternatively, these findings may highlight a disconnect in the sampled voxels along the path of SLF by the two techniques (DTI and 1H-MRS) due to image misalignment and differences in spatial resolutions. Therefore, there is an unmet need to develop a user-independent, objective and comprehensive template-based data analytical method for simultaneously analyzing metabolite and microstructural changes from similar voxels along the path of SLF in several neurological disorders.

The whole brain spectroscopic imaging (WBSI) provides high-resolution metabolic maps spanning both supratentorial and infratentorial regions(Ebel, Soher, & Maudsley, 2001; A. Maudsley et al., 2006). These maps can be spatially co-registered to anatomical images, thus facilitating mapping of metabolite alterations from multiple brain regions(Chawla et al., 2015; Verma et al., 2019). Some studies have demonstrated WBSI’s potential to detect widespread metabolite alterations from multiple brain regions in amyotrophic lateral sclerosis (ALS) (Govind et al., 2012; Verma et al., 2013) and multiple sclerosis (MS) (Donadieu et al., 2016). On the other hand, neurite orientation dispersion and density imaging (NODDI) is an advanced and more sensitive dMRI technique for detecting microstructural damage than conventional diffusion tensor imaging (DTI) sequence(Caverzasi et al., 2016; Zhang, Schneider, Wheeler-Kingshott, & Alexander, 2012). Tractometry is an advanced neuroimaging technique used to analyze the microstructural properties of white matter tracts in the brain(Bells et al., 2011; Yeatman, Dougherty, Myall, Wandell, & Feldman, 2012). Unlike traditional voxel-based or region of interest (ROI) approaches that provide average values across a defined region, tractometry quantifies parameters along the entire length of specific fiber tracts.

With unmet need of integrating pixel-by-pixel WBSI and NODDI/DTI derived parametric maps along the path of SLF in mind, this proof-of-concept study was designed to (i) fuse WBSI and NODDI/DTI derived parametric maps along the path of SLF I and II segments and quantitatively assess the spatial concordance (ii) evaluate regional metabolite concentrations and microstructural properties along the trajectories of the SLF I and II segments and (iii) determine the relationships among WBSI and NODDI/DTI derived parameters from SLF I and II segments in normal healthy adults. Our central hypothesis was that simultaneous analysis of metabolic and microstructural profiles from anatomically distinct segments of SLF will reveal new insights into the complex pathophysiological processes occurring along its path. Once established in normal healthy adults, this novel methodology can be applied in patient populations with various pathological conditions in future research and clinical studies.

## Methods

### 1. Data Acquisition

#### 1.1 Healthy Adult Individuals

This study was approved by the Institutional Review Board and was compliant with the HIPAA (the Health Insurance Portability and Accountability Act). A cohort of 10 normal healthy adult individuals (4F, 6M; Age: 31.4 ± 7.53 years) were recruited for this study. The criteria for including normal healthy individuals were as follows: (i) between the ages of 18-45 years (ii) physically healthy with a normal medical record/history (iii) willing to undergo MRI. Criteria for excluding individuals are as follows: (i) pregnancy (ii) lactation (iv) history of type-2 diabetes, hypoglycemia, respiratory, cardiovascular or neurological/ neuropsychological disorders, diseases (i.e. heart attack, stroke) as reported in medical records (vii) claustrophobia (viii) harboring any MRI contraindicated biomedical device/implant. All individuals were screened by two physicians (investigators on this study) based on medical records/history, physical examination, and vital signs before considering them as healthy individuals.

All healthy individuals underwent an MRI on the same scanner with identical protocol only at one timepoint. However, a single individual (about 30-35 years old) underwent MRI three times with scans spaced roughly a week apart to examine the intrasubject variability of tractometry profiles.

### 1.2 MR Imaging

All MRI data were acquired on a Siemens 3T Magnetom PrismaFit (Erlangen, Germany) scanner equipped with a 64-channel phased array coil. The MRI protocol included a three plane localizer, an axial three-dimensional (3D)-T1-MPRAGE (magnetization-prepared rapid acquisition of gradient echo) imaging with the following parameters: repetition time (TR)/ echo time (TE)/inversion time (TI) = 1620/3.9/950 ms; matrix size = 192 × 256; section thickness = 1 mm; number of sections per slab = 192; flip angle = 15°; number of excitations (NEX) = 1; bandwidth (BW) = 150 Hz/pixel and axial T2-fluid attenuated inversion recovery (T2-FLAIR) imaging (TR/TE/TI = 9420/141/2500 ms, slice thickness = 3 mm; the number of slices = 60).

#### 1.2.1 Diffusion MRI (dMRI)

A multi-shell, high angular resolution diffusion imaging (HARDI) protocol with 3 b-values and 109 unique non-collinear diffusion encoding directions was used for this study. The protocol consisted of b-values of 300, 800, and 2000 s/mm^2^, with a 2mm isotropic voxel size. Nine interspersed non-diffusion encoded b_0_ images were also acquired, resulting in a total of 118 volumes. Other scan parameters were: a TR/TE = 4420/78.8 ms, matrix size = 110 x 110, flip angle = 90°, number of slices = 76, and multiband acceleration factor of 2. The same protocol was acquired twice with the opposite phase encoding directions [anterior-Posterior (AP) and Posterior-Anterior (PA)] to correct for susceptibility/geometric induced image distortions. The total scan time was about 10 minutes in a single-phase encoding direction.

#### 1.2.2 Whole Brain Spectroscopic Imaging (WBSI)

3D-WBSI data were acquired for the whole brain including supratentorial and infratentorial regions except for portions of the anterior frontal cortex, brainstem, and cerebellum. A single 50mm thick outer volume saturation band was positioned to cover the skull base and the paranasal sinuses. An inversion recovery-based lipid nulling sequence (TI=198ms) was also used to minimize lipid signal contamination from the skull and scalp outside the brain. A Chemical shift selective saturation (CHESS) based sequence was also used for water suppression. The data were acquired in an interleaved manner, with a modified spin-echo sequence for the acquisition of metabolites, and a gradient-recalled echo sequence for water signal acquisitions, with parallel imaging using generalized auto-calibrating partially parallel acquisition (GRAPPA) and an acceleration factor of 1.6 (A. Maudsley et al., 2006). Acquisition parameters included: TR/TE = 1550/17.6 ms, NEX = 1, field of view = 280 × 280 × 180 mm^3^, matrix size = 50 × 50 × 18, voxel size = 5.6 × 5.6 × 10 mm^3^ (≈0.31 cm^3^) spectral points = 512, BW = 625 Hz, and excitation angle = 73°. The total acquisition time was 17 min 50 s. To ensure high-quality data, manual shimming was performed to achieve an optimal fullwidth at half-maximum value (<25 Hz) of the magnitude water signal before every acquisition.

### 2. Data Processing

The overall image processing pipeline to reconstruct the microstructural and metabolite tract profiles for SLF I-II segments is illustrated in **Figure 1**.

**Figure 1.**
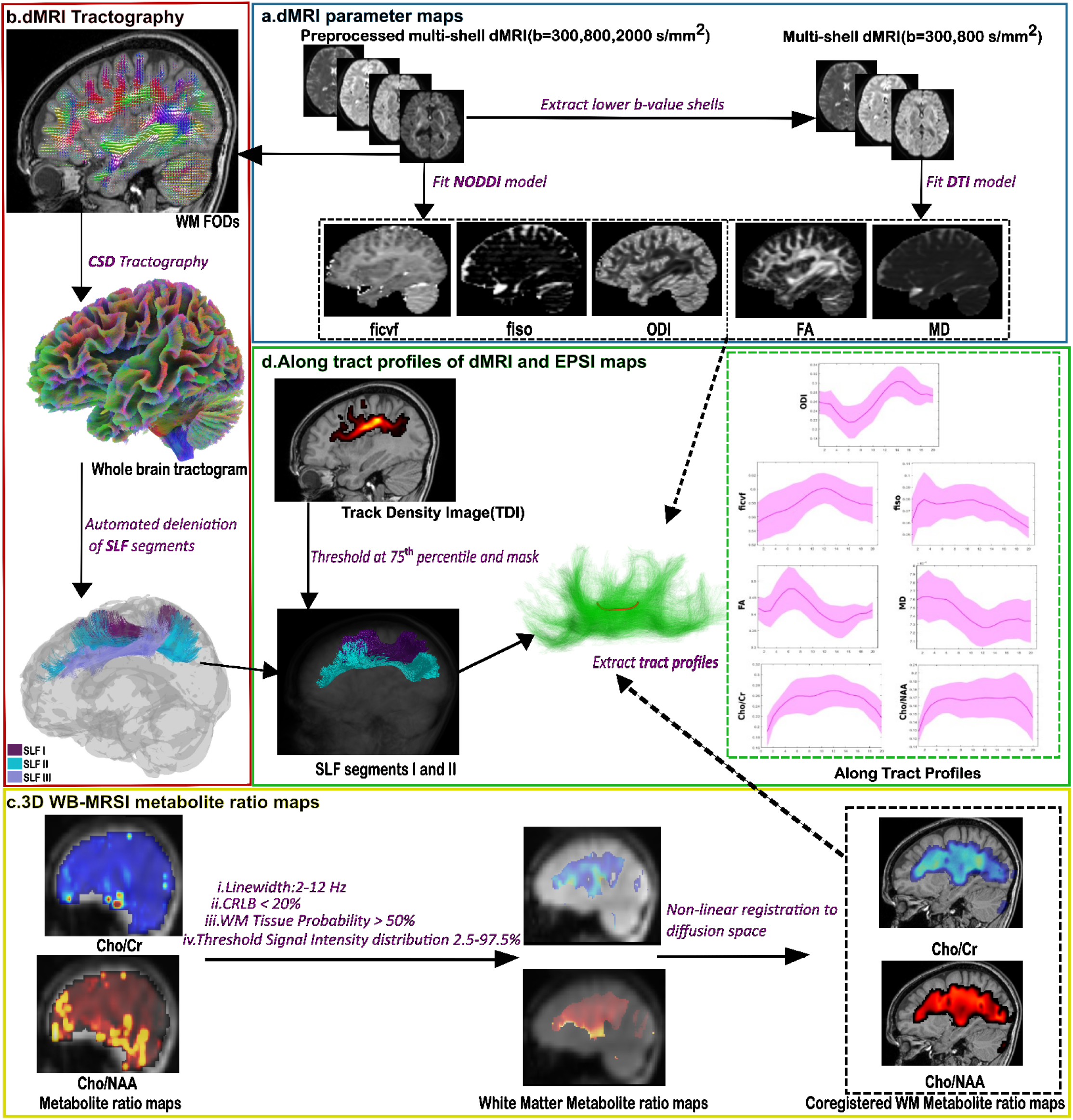
The overall image processing pipeline to reconstruct tract profiles for SLF I-II segments. The multi-shell HARDI data were used to generate DTI and NODDI derived maps (a) Constrained spherical deconvolution (CSD) tractography was performed for automatic generation of tracts (b) WBSI maps after assessment of spectral quality and inclusion of only white matter tissues were co-registered to the diffusion space (c) Tract profiles were finally generated for the NODD/DTI and WBSI-derived parameters along core tract centroids divided into 20 distinct sections (d).

### 2.1 dMRI preprocessing, tractography, and generation of DTI and NODDI metrics

The dMRI data preprocessing was performed using the FSL (S. M. Smith et al., 2004) and MRtrix3 (J.-D. Tournier et al., 2019) software. Initially, the raw DICOM data files were converted to the NIfTI format. The two sets of images acquired with opposite phase encoding directions were combined, creating a single dataset prior to applying denoising algorithm (Veraart et al., 2016) using Marchenko-Pastur principal component analysis (MP-PCA). Subsequently, Gibbs ringing artifact correction was performed using the method of sub-voxel shifts (Kellner, Dhital, Kiselev, & Reisert, 2016). Further, susceptibility distortion and eddy current induced artefacts were corrected using FSL’s *eddy* (Andersson & Sotiropoulos, 2016) and *topup* (Andersson, Skare, & Ashburner, 2003) tools. The resulting data were subjected to bias field correction (Tustison et al., 2010). The b_0_ volumes were extracted from the data and averaged to create a single b_0_ image. Skull-stripping was performed on the mean b_0_ volume (S. M. Smith, 2002) and the corresponding brain mask was used in the subsequent steps to limit the data computation within the voxels of the brain.

The tissue response functions for the white matter (WM), grey matter (GM), and cerebrospinal fluid (CSF) were estimated for each shell using an unsupervised algorithm (Dhollander, Raffelt, & Connelly, 2016). A multi-shell, multi-tissue spherical deconvolution algorithm was used to generate the fiber orientation dispersion function (FOD) for the three tissue types (WM, GM and CSF) (Jeurissen, Tournier, Dhollander, Connelly, & Sijbers, 2014). A global intensity normalization of the FODs was also performed (Raffelt et al., 2017). The WM FODs were used to construct whole brain tractograms using a probabilistic tracking algorithm (J. D. Tournier, Calamante, & Connelly, 2010), anatomically constrained (R. E. Smith, Tournier, Calamante, & Connelly, 2012) by the white matter segmentations on the T1 MPRAGE image coregistered to the b_0_. The three segments of the SLF were generated from the WM FODs using an automated tool (*TractSeg*) that performs a probabilistic bundle tracking of 72 major white matter tracts using a U-Net-based model trained on the WM FOD peaks (Wasserthal, Neher, & Maier-Hein, 2018). The default parameters were used for the FOD cut-off, minimum length of streamlines, step size, and maximum angle. The number of streamlines was set to 8000 for each tract. For each subject, the anatomical congruence of the generated SLF segments I and II was manually verified by two neuroradiologists (L.L.G and M.H).

The preprocessed dMRI data were also used to compute the DTI and NODDI-derived parametric maps. Only the first two lower b-valued shells (b=300 s/mm^2^ and b=800 s/mm^2^) were used to generate mean diffusivity (MD) and fractional anisotropy (FA) maps by applying a voxel-wise weighted least squares fit of a diffusion tensor model (Basser, Mattiello, & LeBihan, 1994) to the preprocessed data implemented in *dwi2tensor* and *tensor2metric* commands. The entire preprocessed data including the higher b-value shell (b=2000 s/mm^2^) was used to fit the NODDI model using a MATLAB (version R2023a, MathWorks, Natick, Massachusetts) based toolbox (Zhang et al., 2012) to derive the indices of free-water fraction (FWF)/isotropic volume fraction (fiso), neurite density index (NDI)/intracellular volume fraction(ficvf), and the orientation dispersion index (ODI). The parameter ficvf quantifies the density of neurites (axons and dendrites) and offers a more precise assessment of neural tissue integrity than DTI derived FA.

**Figure 2.**
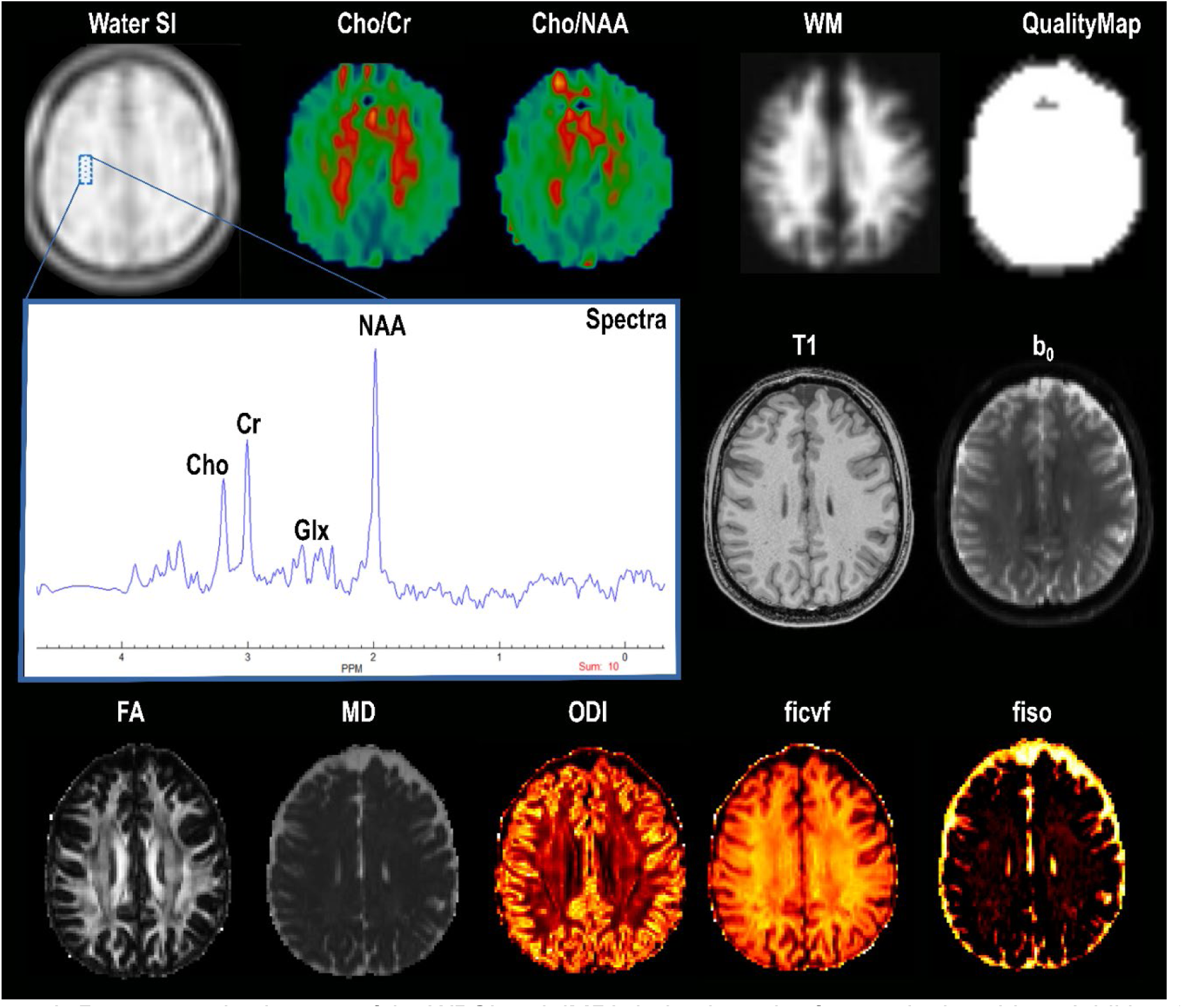
Representative images of the WBSI and dMRI-derived metrics from a single subject. Additionally, the anatomical images, The water reference signal intensity map (WaterSI), the WM tissue probability map (WM), the spectral quality map, and a voxel averaged spectra from 10 white matter voxels are shown. Cho: Choline. Cr: Creatine. Glx: Glutamate + Glutamine. NAA: N-Acetyl Aspartate. FA: Fractional Anisotropy. MD: Mean Diffusivity. ODI: Orientation Dispersion Index. ficvf: Intra-cellular volume fraction. fiso: isotropic volume fraction.

Isotropic volume fraction (fiso) measures the fraction of free water diffusion, allowing for the separation of free water and tissue compartments. This is particularly useful in identifying areas of edema or CSF contamination. ODI assesses the variability in neurite orientation, providing insights into the complexity of neural architecture especially in regions with crossing or intersecting fibers. To minimize the impact of partial volume averaging from CSF, the ficvf and ODI maps were modulated by the tissue fraction maps.

### 2.2 WBSI Preprocessing

The WBSI data were analyzed using metabolic imaging and data analysis system (MIDAS) (A. Maudsley et al., 2006) package (version 2.35) based on IDL version 8.8.2 (Exelis Visual Information Solutions, Boulder, CO, USA). The acquired data, with an initial matrix of 50 × 50 × 18, were zero-filled to a final matrix size of 64 × 64 × 32, yielding an interpolated voxel size of 4.3 × 4.3 × 5.6 mm^3^ (≈0.10 cm^3^). The standard preprocessing steps included field inhomogeneity and eddy current correction, k-space re-gridding, spatial and spectral Fourier transformation, spatial registration with the anatomical T1 image, lipid k-space extrapolation and signal normalization(A. Maudsley et al., 2006; Verma et al., 2019). In each case, quality assurance was evaluated by considering Cramer-Rao lower bounds (<20%), line shape, line width (2-12Hz), CSF contamination, and degree of residual water and lipid signals. The parametric maps of choline/N-acetylaspartate (Cho/NAA) and choline/creatine (Cho/Cr) were computed within the MNI space with a 2mm isotropic voxel resolution.

Subsequently, only those voxels that had a greater than 50% probability of white matter tissue were retained for further analysis (Govind et al., 2012; Li et al., 2022). Finally, the voxels with potentially affected signal intensities (either too high or too low) were discarded by thresholding the signal intensities below 2.75% and 97.5% levels of the total signal intensity distribution as reported earlier (Li et al., 2022). The thresholded white matter metabolite maps of Cho/NAA and Cho/Cr were used for further analysis. The WBSI, DTI, and NODDI derived parametric maps from a representative subject are shown in **Figure 2**.

### 2.3 Assessment of Co-localization of dMRI and WBSI images

The co-localization of metabolite maps to dMRI data was acheived using a non-linear registration process. This process utilized the “WaterSI” (Water Signal Intensity) reference image in MNI (Montreal Neurological Institute) space to align the metabolite maps to the dMRI-derived maps, allowing for comparison and analysis of metabolic and diffusion parameters together (See sections 2.1 and 2.2). The WaterSI images were chosen for coregistrations as they exhibit superior image contrast to the metabolite maps. Given that WaterSI images and metabolite maps are inherently registered to each other, the same non-linear transformations could be applied to the metabolite maps. The T1 weighted images were coregistered to the b_0_ images using a rigid-body registration, following which, the WaterSI reference images were non-linearly registered to these T1 weighted images. The same non-linear transformations and warps were finally applied to the preprocessed white matter metabolite images, so as to spatially align them to the b_0_ image. Such an approach was performed since the T1 and WatesSI images showed a similar gray matter/white matter contrast. A few slices of the coregistered T1, coregistered WaterSI and the b_0_ image for a single subject are shown in **Suppementary Figure S1**. To quantitatively asess the degree of co-localization, slice-wise mutual information between these coregistered images and the Pearson’s correlation coefficients were also computed (**Figure 3** and **Table 1**).

### 2.4 Tractometry: generation of tract profiles

The final step after the white matter microstructural and metabolite maps are aligned in the diffusion space, and the tractography of the SLF I and II segments performed, would be to project/sample these values along the length of the tract. Tractometry is a multistep process that includes extracting bundles from a whole brain tractogram, finding a descriptive centre pathway for each bundle, dividing the bundles into equidistant sections along the bundle, and mapping quantitative features along bundles of interest to create a *tract profile*. Bundle segmentation ensures precise delineation of white matter tracts by reducing variability and eliminating outliers. Streamline ordering standardizes directional coherence within and across subjects, facilitating consistent anatomical comparisons. Core streamline definition generates a representative trajectory by resampling streamlines, improving spatial alignment and accounting for variations in length and orientation. Finally, measure assignment maps dMRI-derived metrics (or other metrics co-registered to diffusion space (Dennis et al., 2018; Schiavi et al., 2022)) along the core streamline using robust distance metrics, enhancing alignment and ensuring accurate profiling for statistical analysis(Chamberland, St-Jean, Jones, Descoteaux, & Leemans, 2025).

The WaterSI image generated in the MNI space was coregistered with the T1 weighted image in diffusion space (See section 2.3), using a symmetric normalization algorithm implemented in the ANTs software (Advanced Normalization tools; (Avants, Tustison, & Song, 2009)) and the transformations applied to the thresholded (See section 2.2 on WBSI processing) Cho/NAA and Cho/Cr white matter maps for each tract.

The binarized masks for the metabolite maps in the b_0_ space, and the 75% thresholded track density images (TDI) for each segment of the SLF were used to further constrain the anatomy of the tract to its core for both the WBSI derived Cho/Cr, and Cho/NAA, NODDI derived ficvf, fiso, ODI and DTI derived MD and FA parameters. The estimation of tract profiles was implemented in the *Dipy* (Garyfallidis et al., 2014) and *scilpy* (Renauld et al., 2026) software. It involved calculating the centroid of each tract, orienting the streamlines along the tract centroid, and projecting the values along the tract by uniformly dividing it into 20 discrete but spatially contiguous sections (Chamberland et al., 2019). This process resulted in a total of 7 distinct tract profiles (5 dMRI and 2 WBSI) for the SLF segments I and II bilaterally. Subsequently, section-wise mean values of MD, FA, fiso, ficvf, ODI, Cho/NAA, and Cho/Cr tract profiles were computed from all subjects. The two end sections were removed from the tract profiles before any further analysis to reduce anatomical variability which is generally found prominently along the terminal regions of the tract.

### 2.5 Intra-Subject Analysis

To test the reliability of tract reconstruction and the repeatability of microstructure and metabolite derived tract profiles, a single individual underwent MRI three times with the scans spaced roughly a week apart.

Specifically for the intra-subject analysis, a single subject derived b_0_ template was generated from the preprocessed b_0_ images of all the three time points. The *antsMultivariateTemplateConstruction2.sh* script implemented in ANTs was used to accomplish this. For each of the time points, the final tractography outputs, the dMRI derived maps, and the WBSI derived maps in their individual diffusion space were transformed to the common b_0_ template prior to performing tractometry, and further analyses.

The reliability of reconstructed tracts was assessed using the Dice similarity coefficients (an index of spatial mask overlap between two data sets), the repeatability was assessed using Bland-Altman plots, coefficient of variation (CV), and repeatability coefficient (RC)

#### 2.5.1 Dice coefficients

To verify the reliability of the tractography algorithm, the overlap of the SLF tracts reconstructed for each pair of time points were assessed using a standard Dice-similarity coefficient overlap of the reconstructed tracts aligned to the single subject b_0_ template. However, since these white matter tracts have more dense and coherent streamlines towards the center than at the periphery, a tract density weighted dice-coefficient (Cousineau et al., 2017) was also calculated between each pair of acquisitions, to penalize for spurious streamlines located further away from the tract core(**Table 2** and **Figure 4**). Values greater than 70% were considered acceptable.

#### 2.5.2 Repeatability Analysis

For the single-subject data, repeatability analysis was also carried out for each tract profiles for the bilateral SLF segments I and II acquired at three time points. The Bland-Altman plots were calculated for each pair of acquisitions for every parameter across the 20 segments (**See supplementary Figure S2**). The RC, and the CV were determined. All the analyses were performed in Python (version 3.9.12) using the *numpy* (Harris et al., 2020)(version 1.22.4) and *pandas* (version 2.2.2) libraries.

### 2.6 Inter-Subject Analysis

The section-wise coefficient of variation for all the 10 subjects was also computed and the mean CVs reported as Inter-subject CV. To investigate the potential linear associations between the WBSI and dMRI tract profiles, single and multiple-variable regression analyses were performed. For each of the 20 segments, the dMRI and WBSI tract profile values were averaged across the 10 participants to generate the mean tract profiles for both WBSI and dMRI metrics, which were subsequently compared. The metabolite tract profiles (Cho/Cr and Cho/NAA) were considered as the dependent variable. The significance level was set at p<0.01. The python libraries *scikit-learn* (Pedregosa et al., 2011) and *statsmodels* (Seabold & Perktold, 2010) were used for all the regression analyses.

### 2.7 Relationships between WBSI and NODDI derived Parameters

To investigate the relationships among WBSI derived parameters (Cho/Cr and Cho/NAA) and NODDI/DTI derived parameters (ficvf, ODI, fiso, FA, and MD), Pearson’s correlation coefficient analyses were performed. A probability (p) value of less than 0.01 was considered significant. Additionally, multivariate logistic regression analyses with backward stepwise selection were used to ascertain the relationship between multiple independent parameters.

## 3. Results

In general, good quality WBSI data was obtained from all subjects. Quality map analysis demonstrated that a significant percentage (∼ 70%) of the brain voxels within each subject’s data had good quality. The majority of voxels with insufficient quality suffered from a broad spectral linewidth due to field distortions caused by poor B_0_ homogeneity, magnetic susceptibility artifacts and lipid contamination. The poor-quality spectra were mainly observed from brain stem regions.

### 3.1 Co-localization of dMRI and WBSI derived Images

**Figure 3.**
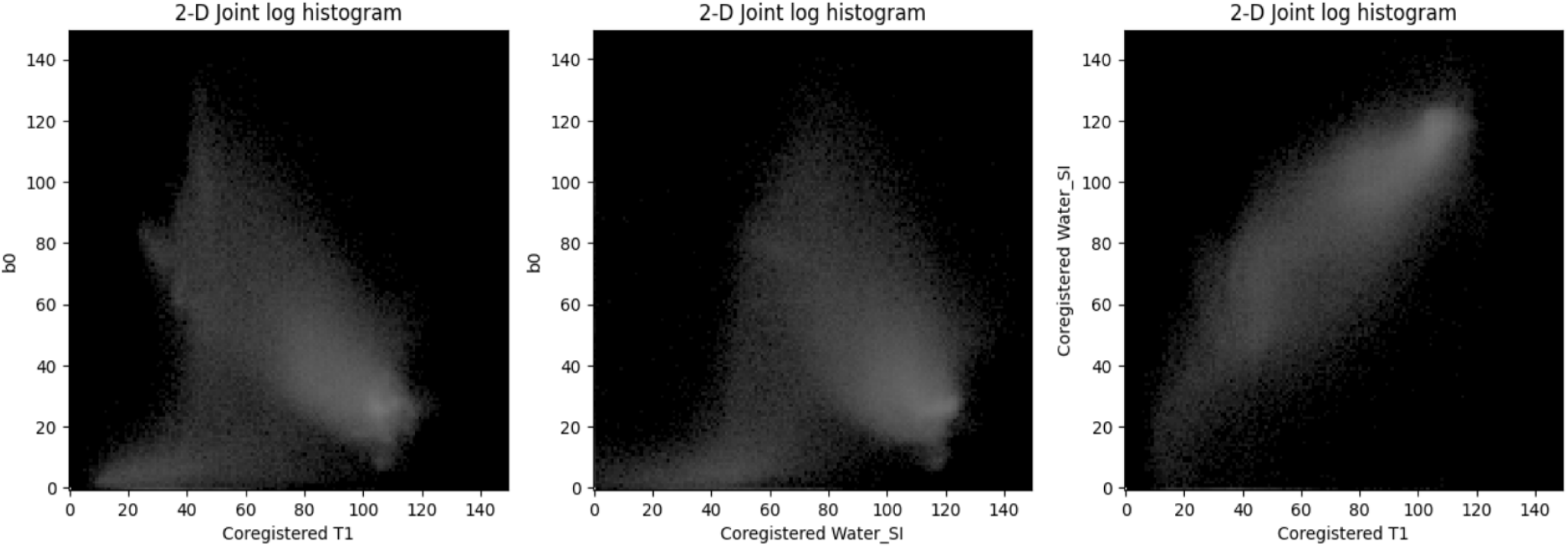
2D-Joint log-transformed intensity histogram plots of co-registered images in the diffusion (b_0_) space from a representative subject. The grayscale represents natural logarithm of paired pixel frequency.

The 2D log histograms of pairwise images from a representative subject are shown in **Figure 3**. These histograms show a strong and direct relationship between the coregistered T1 and WaterSI images reflecting their similar GM-WM contrast on both image types. Conversely, the coregistered b_0_ images show strong but inverse relationships with both T1 and WaterSI images, highlighting their dissimilar GM-WM contrast.

The mutual information (MI) from all subjects is presented in **Table 1**. On visual inspection, a good concordance between the dMRI and WBSI images was observed from all subjects (**Supplementary Figure S1**). This initial observation was later confirmed by a quantitative analysis using mutual information between the co-registered images in all subjects (**Table 1**). The mean (±SD) values for MI and normalized MI (NMI) between T1 weighted and b_0_ images were 1.005 ± 0.076 and 0.584 ± 0.0156 respectively with an average Pearson’s correlation (r) of 0.836 ± 0.037. The co-registered b_0_ and WaterSI image had an MI= 0.9513 ± 0.046; NMI= 0.572 ± 0.019; and r= 0.882 ± 0.029.

**Table 1.** Mutual information (MI), normalized mutual information (NMI), and Pearson’s correlation (r) in all subjects (n=10)

| Subject | MI,NMI,r<br>($b_0$, T1_coreg) | MI, NMI,r<br>($b_0$, Water_SI_coreg) | MI, NMI,r<br>(T1_coreg, Water_SI_coreg) |
| --- | --- | --- | --- |
| SUB_001 | 0.928,0.599,0.872 | 0.890,0.586,0.912 | 0.931,0.600,0.986 |
| SUB_002 | 1.008,0.583,0.820 | 0.962,0.570,0.875 | 1.008, 0.587, 0.984 |
| SUB_003 | 0.931,0.586,0.863 | 0.907,0.579,0.902 | 0.932, 0.591, 0.983 |
| SUB_004 | 1.030,0.587,0.813 | 0.994,0.576,0.865 | 1.023, 0.587, 0.987 |
| SUB_005 | 1.029, 0.578, 0.852 | 0.988, 0.566, 0.892 | 1.041, 0.587, 0.988 |
| SUB_006 | 0.912, 0.605, 0.845 | 0.885,0.597,0.883 | 0.910, 0.611, 0.987 |
| SUB_007 | 1.041,0.588,0.769 | 1.008, 0.578, 0.821 | 1.048, 0.593, 0.984 |
| SUB_008 | 1.170,0.546,0.785 | 1.010,0.526,0.856 | 1.142, 0.541,0.981 |
| SUB_009 | 0.968,0.581,0.869 | 0.945, 0.574, 0.906 | 0.962, 0.586, 0.989 |
| SUB_010 | 1.033,0.586, 0.869 | 0.974, 0.566, 0.907 | 1.006, 0.580, 0.986 |

**Figure 4.**
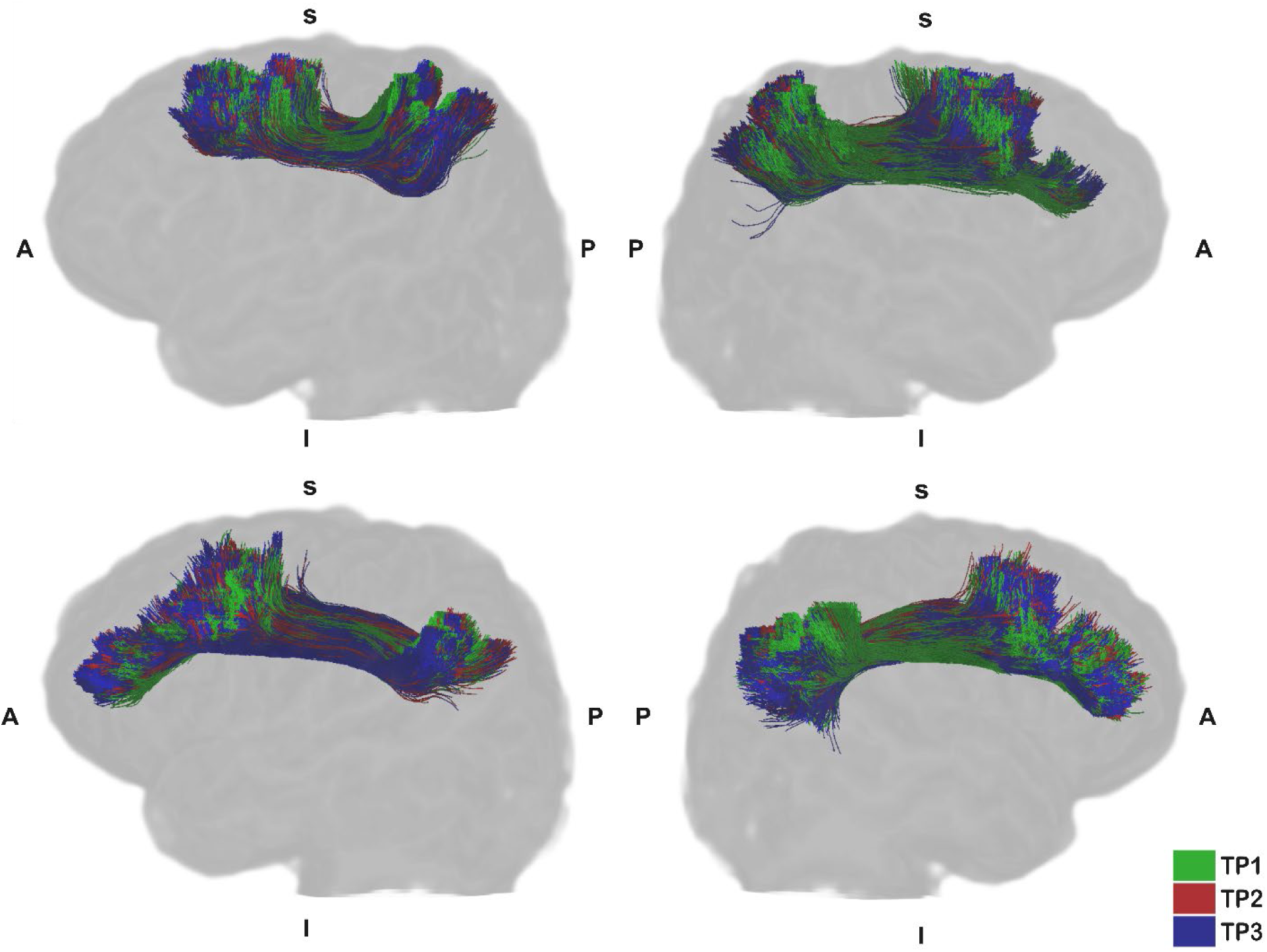
The SLF I and II reconstructed from dMRI data for the same subject scanned at 3 different time points (TP1, TP2, TP3) were co-registered to a single subject derived b_0_ template generated from the individual b_0_ images at three time points. A-Anterior; P-Posterior; S-Superior; I-Inferior

**Table 2:**
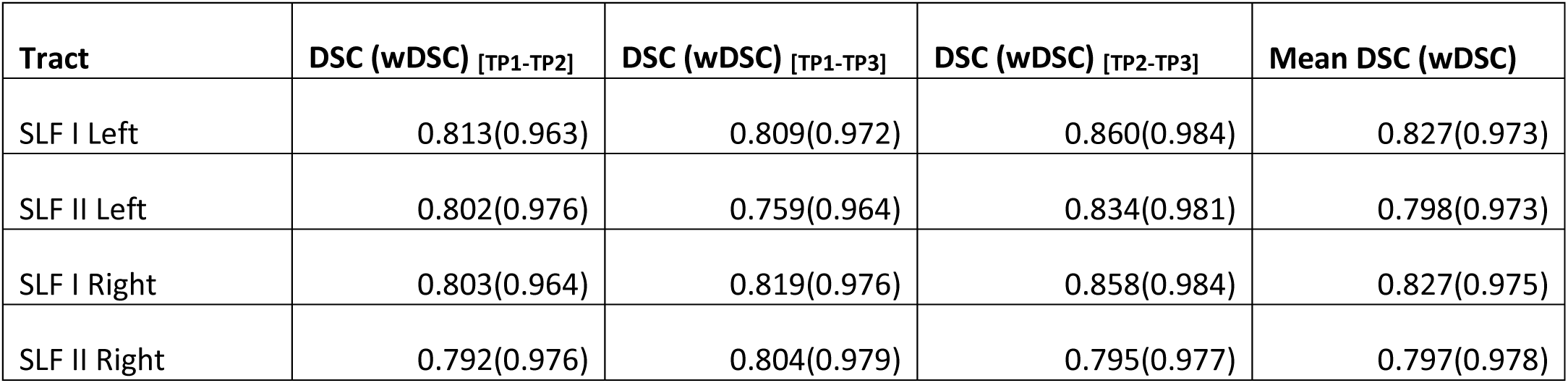
Dice similarity coefficients (DSCs) and track density-weighted Dice coefficients(wDSCs) for bilateral SLF I and II from a single subject scanned at 3 different time points (TP1, TP2, TP3)

The co-registered WaterSI and T1 weighted had an MI = 1.0003 ± 0.070; NMI = 0.586 ± 0.018; and r = 0.986 ± 0.002. The histogram plots suggested a linear relationship between the coregistered T1 and WaterSI images.

### 3.2 Intra and Inter-subject variability in tract profiles

The Bland Altman plots revealed that all the tracts in general had narrow ranges between upper and lower levels of agreement in the section-wise values across acquisitions (**Supplementary Figure S3**). As summarized in **Table 2**, both the standard (DSC) and the track density weighted (wDSC) dice coefficients were greater than 70%, indicating substantial level of agreement.

The results of the repeatability analysis for tract profiles are presented in **Table 3**. The dMRI parameters exhibited higher repeatability, indicating greater consistency in repeated measurements due to RC, and lower intra-subject CV than WBSI derived parameters. Among all dMRI parameters, ficvf was found to show the best repeatability from tract profiles. In general, WBSI derived Cho/Cr showed better repeatability than Cho/NAA from tract profiles. Additionally, the SLF I right showed the best repeatability across all measured parameters in comparison with other tracts. The inter-subject CVs, while generally higher than the intra-subject CVs reflecting higher inter-individual variability in tract profiles, were ∼3-17% for all the measures except the free water fraction (∼18-23%).

**Table 3.** Repeatability analysis of tract profiles.

| **Measure** | **RC**<br>**LOA: [RC, -RC]** | **CV%**<br>**(Intra-subject)** | **CV%**<br>**(Inter-subject)** |
| --- | --- | --- | --- |
| **SLF I Right** |  |  |  |
| **ODI** | 0.258 | 2.545 | 12.27 |
| **ficvf** | 0.183 | 0.830 | 4.43 |
| **fiso** | 0.116 | 1.675 | 20.5 |
| **FA** | 0.378 | 2.671 | 9.95 |
| **MD** | 0.00014886 | 0.666 | 3.50 |
| **Cho/Cr** | 0.446 | 5.641 | 12.51 |
| **Cho/NAA** | 0.296 | 6.837 | 14.42 |
| **SLF I Left** |  |  |  |
| **ODI** | 0.259 | 0.702 | 11.27 |
| **ficvf** | 0.182 | 0.415 | 4.57 |
| **fiso** | 0.139 | 9.482 | 21.98 |
| **FA** | 0.434 | 0.689 | 8.90 |
| **MD** | 0.0001309 | 0.707 | 3.19 |
| **Cho/Cr** | 0.444 | 5.321 | 9.78 |
| **Cho/NAA** | 0.281 | 9.314 | 12.51 |
| **SLF II Left** |  |  |  |
| **ODI** | 0.307 | 1.241 | 12.72 |
| **ficvf** | 0.119 | 0.756 | 4.86 |
| **fiso** | 0.123 | 5.663 | 23.10 |
| **FA** | 0.348 | 2.055 | 12.10 |
| **MD** | 0.00007458 | 1.036 | 3.28 |
| **Cho/Cr** | 0.415 | 0.618 | 13.53 |
| **Cho/NAA** | 0.269 | 2.469 | 17.55 |
| **SLF II Right** |  |  |  |
| **ODI** | 0.389 | 2.007 | 11.61 |
| **ficvf** | 0.138 | 1.071 | 4.55 |
| **fiso** | 0.118 | 5.162 | 18.21 |
| **FA** | 0.524 | 1.846 | 11.13 |
| **MD** | 0.000077 | 0.245 | 3.28 |
| **Cho/Cr** | 0.525 | 1.403 | 11.14 |
| **Cho/NAA** | 0.352 | 3.934 | 13.89 |

**Figure 5.**
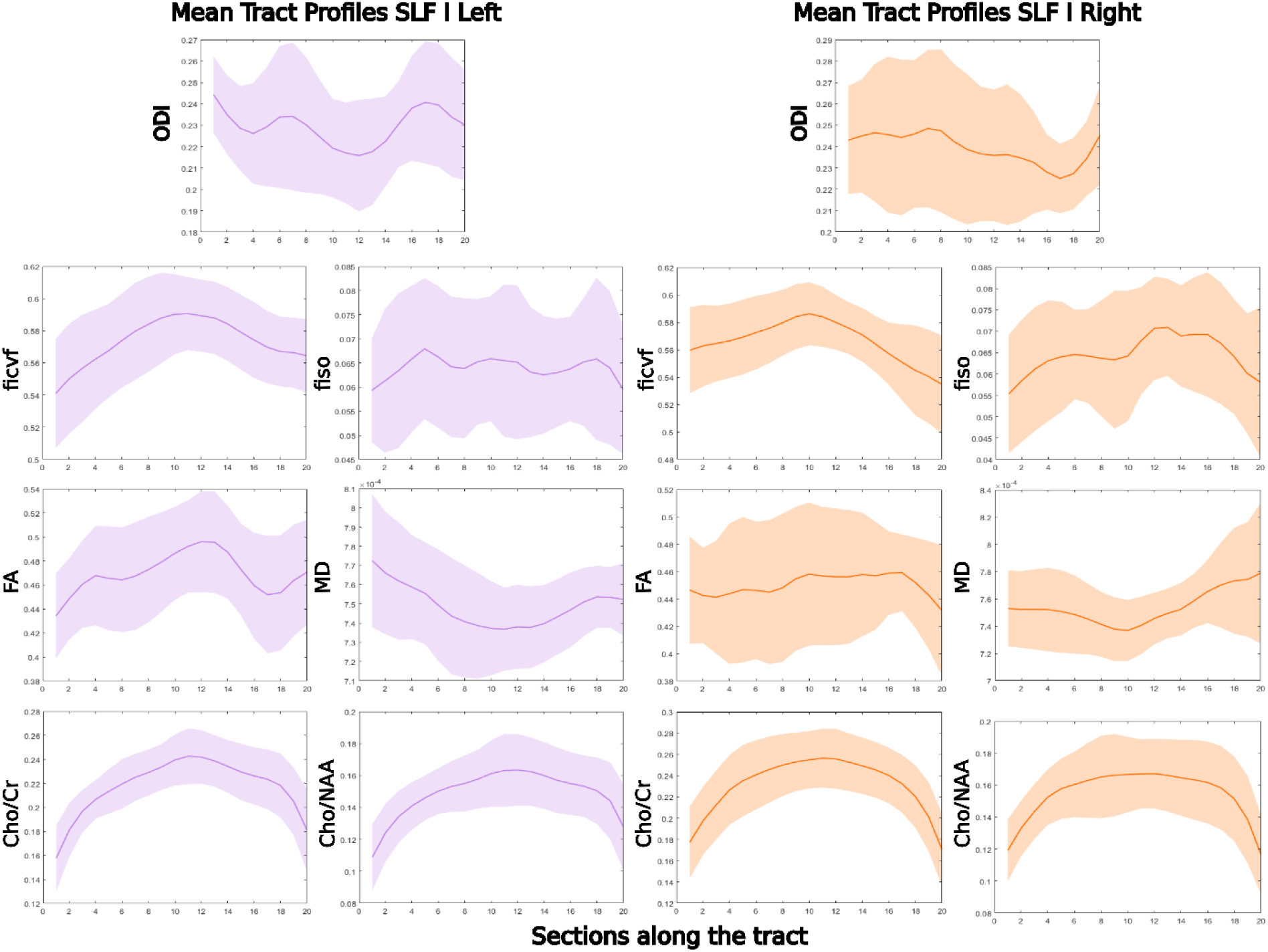
Mean Tract profiles of WBSI and NODDI/DTI derived parameters in the bilateral SLF I from 10 healthy subjects

**Figure 6.**
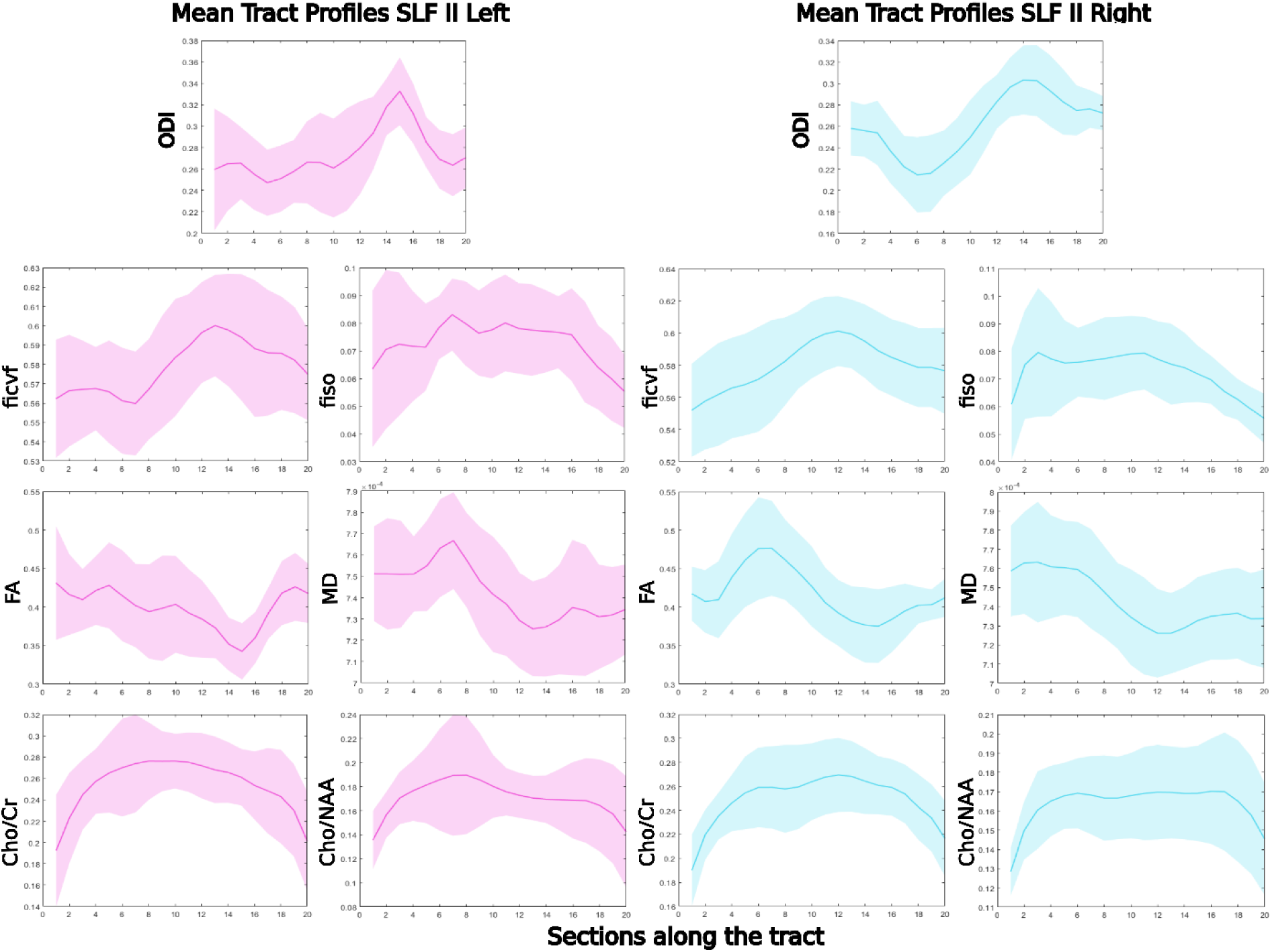
Mean Tract profiles of WBSI and NODDI/DTI derived parameters in the bilateral SLF II from 10 healthy subjects

### 3.3 Associations between dMRI and WBSI derived tract profiles

The mean (averaged across 10 subjects for each of the 20 segments per tract) tract profiles from 10 subjects are shown in **Figure 5** and **Figure 6**. Single variable regression analyses revealed significant linear associations between the dMRI and WBSI tract profiles. Cho/Cr tract profile showed positive and significant (p<0.01) linear associations with ficvf, and FA, and a negative and significant (p<0.01) linear association with MD from bilateral SLF I. Cho/Cr and fiso also had a positive linear association from the right SLF I. In SLF II, Cho/Cr had positive associations with ficvf and fiso and a trend towards negative association with the MD (p>0.01) (**Figure 7**).

Cho/NAA showed comparable associations with dMRI derived parameters as those observed with Cho/Cr from bilateral SLF (**Figure 8**).

As summarized in **Table 4**, when multivariate logistic regression analyses were performed to ascertain the relationship between Cho/Cr and NODDI/DTI derived parameters, the best model was achieved with two parameters (ficvf and MD) as follows.Similarly, when relationship between Cho/NAA and NODDI/DTI derived parameters was tested (**Table 5**.), the best model was achieved with two parameters (ficvf and MD) as follows

**Figure 7.**
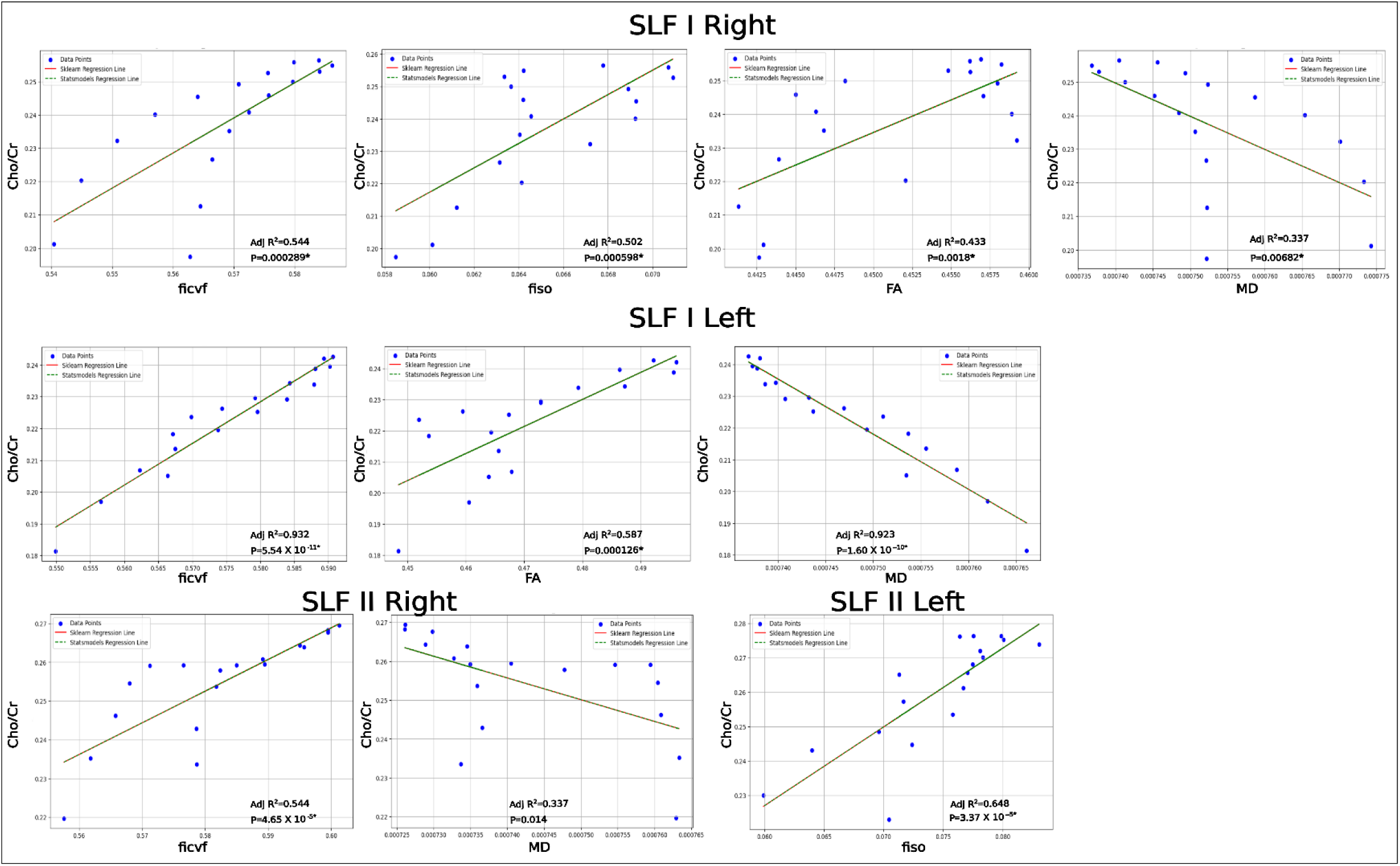
Single variable regressions of the bilateral SLF tract profiles. Cho/Cr vs NODDI/DTI derived parameters. *Significant at p<0.01.

**Figure 8.**
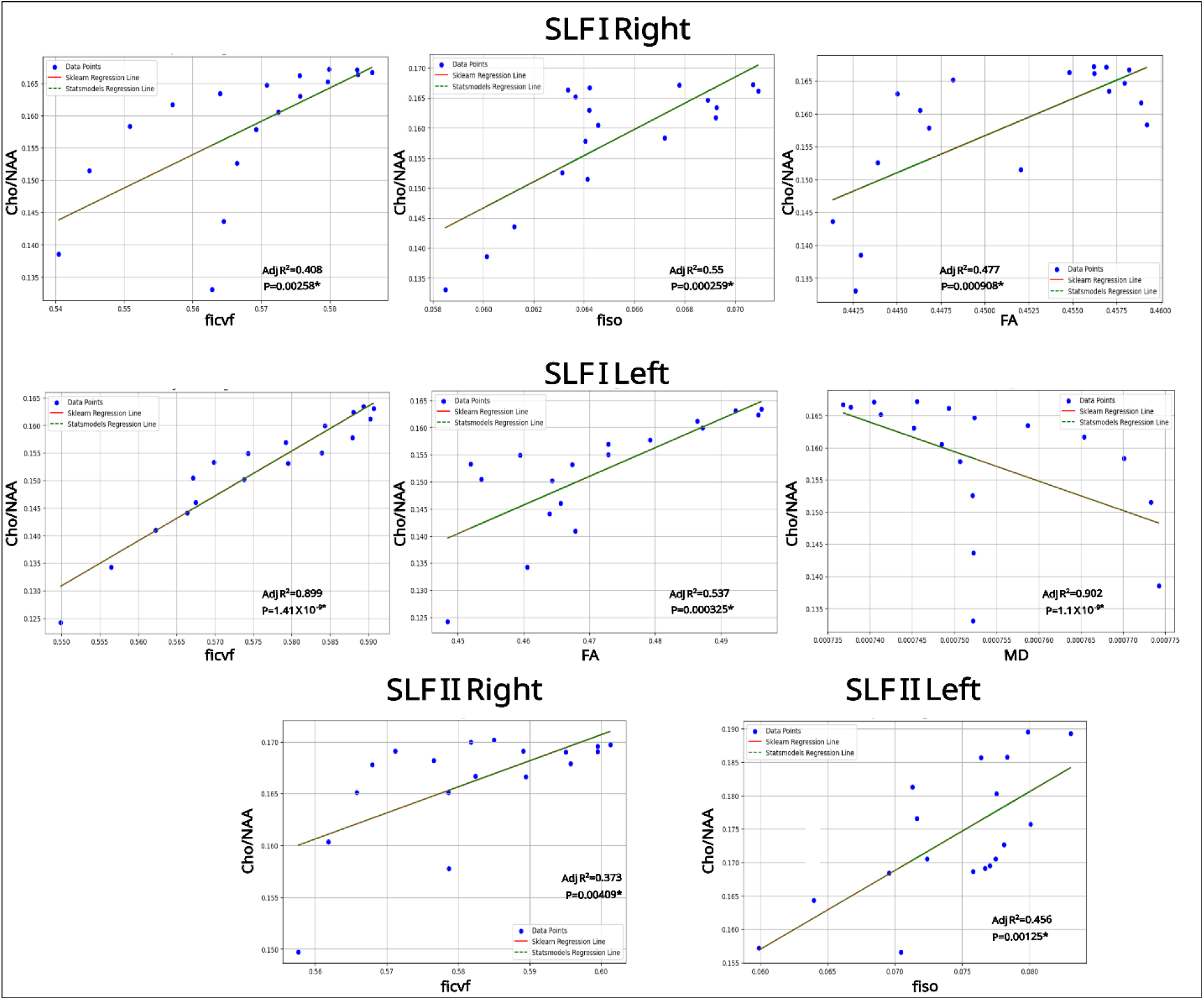
Single variable regressions of the bilateral SLF tract profiles. Cho/NAA vs NODDI/DTI derived parameters. *Significant at p<0.01.

**Table 4:** Multiple variable regression analyses of Cho/Cr with dMRI derived parameters.

| Tract | Adj R<sup>2</sup> | Fstat | Prob(Fstat) | $\beta_0$ | $\beta_1(\text{ODI})$ | $\beta_2(\text{ficvf})$ | $\beta_3(\text{fiso})$ | $\beta_4(\text{FA})$ | $\beta_5(\text{MD})$ |
| --- | --- | --- | --- | --- | --- | --- | --- | --- | --- |
| SLF I right | 0.968 | 103.1 | 1.97E-09 | -18.1979 | 1.9292 | 12.6898 | -5.4852 | 1.1562 | 14090 |
| SLF II right | 0.965 | 96.13 | 2.97E-09 | -10.9855 | -0.1743 | 8.9575 | -5.8796 | -0.7601 | 9186.316 |
| SLF II left | 0.804 | 14.94 | 8.53E-05 | -4.7144 | -1.4055 | 4.5317 | -2.6651 | -1.4214 | 4708.523 |
| SLF I left | 0.945 | 59.2 | 4.87E-08 | 7.2268 | -0.4195 | -3.8957 | 4.1491 | 0.0112 | -6608.323 |
| Tract | Adj R<sup>2</sup> | Fstat | Prob(Fstat) | $\beta_0$ | $\beta_1(\text{ODI})$ | $\beta_2(\text{ficvf})$ | $\beta_3(\text{fiso})$ | $\beta_4(\text{MD})$ | |
| SLF I right | 0.962 | 108.6 | 7.51E-10 | -18.6516 | 0.937 | 13.7969 | -5.7623 | 1.49E+04 |  |
| SLF II right | 0.955 | 91.03 | 2.27E-09 | -7.2997 | 0.4489 | 5.7955 | -3.2318 | 5792.24 |  |
| SLF II left | 0.794 | 17.37 | 3.94E-05 | -7.1118 | -0.1505 | 5.64 | -1.0819 | 5689.667 |  |
| SLF I left | 0.949 | 80.17 | 5.00E-09 | 7.3008 | -0.4392 | -3.9354 | 4.1713 | -6665.66 |  |
| Tract | Adj R<sup>2</sup> | Fstat | Prob(Fstat) | $\beta_0$ | $\beta_1(\text{ficvf})$ | $\beta_2(\text{fiso})$ | $\beta_3(\text{MD})$ | | |
| SLF I right | 0.942 | 92.77 | 1.78E-09 | -12.1727 | 9.4728 | -3.5554 | 9645.55 |  |  |
| SLF II right | 0.847 | 32.26 | 1.52E-06 | -2.8142 | 2.7761 | -1.0965 | 2064.606 |  |  |
| SLF II left | 0.79 | 22.35 | 1.33E-05 | -8.6933 | 6.697 | -1.999 | 7030.576 |  |  |
| SLF I left | 0.948 | 104.1 | 8.23E-10 | 2.797 | -0.9271 | 2.3122 | -2930.25 |  |  |
| Tract | Adj R<sup>2</sup> | Fstat | Prob(Fstat) | $\beta_0$ | $\beta_1(\text{ficvf})$ | $\beta_2(\text{MD})$ | | | |
| SLF I right | 0.923 | 103.4 | 1.68E-09 | -6.0766 | 4.9795 | 4629.655 |  |  |  |
| SLF II right | 0.811 | 37.53 | 1.45E-06 | -1.5265 | 1.7645 | 1014.677 |  |  |  |
| SLF II left | 0.77 | 29.39 | 6.47E-06 | -4.8866 | 3.8146 | 3953.5 |  |  |  |
| SLF I left | 0.928 | 110.2 | 1.07E-09 | -0.3559 | 1.1986 | -149.065 |  |  |  |

**Table 5:** Multiple variable regression analyses of Cho/NAA with dMRI derived parameters.

| Tract | Adj R<sup>2</sup> | Fstat | Prob(Fstat) | β<sub>0</sub> | β<sub>1</sub>(ODI) | β<sub>2</sub>(ficvf) | β<sub>3</sub>(fiso) | β<sub>4</sub>(FA) | β<sub>5</sub>(MD) |
| --- | --- | --- | --- | --- | --- | --- | --- | --- | --- |
| SLF I right | 0.953 | 69.51 | 1.94E-08 | -12.6263 | 1.536 | 8.5687 | -3.9953 | 0.9355 | 9816.1735 |
| SLF II right | 0.818 | 16.32 | 5.46E-05 | -7.2248 | -0.3344 | 5.8883 | -4.4547 | -0.7171 | 6300.1098 |
| SLF II left | 0.811 | 15.57 | 6.92E-05 | -2.3911 | -1.1627 | 2.3534 | -2.607 | -1.2837 | 2994.9303 |
| SLF I left | 0.954 | 71.52 | 1.65E-08 | 6.8162 | -0.0246 | -4.1816 | 3.7374 | 0.2783 | -6187.3012 |
| Tract | Adj R<sup>2</sup> | Fstat | Prob(Fstat) | β<sub>0</sub> | β<sub>1</sub>(ODI) | β<sub>2</sub>(ficvf) | β<sub>3</sub>(fiso) | β<sub>4</sub>(MD) |  |
| SLF I right | 0.939 | 66.33 | 1.62E-08 | -12.9934 | 0.7331 | 9.4646 | -4.2195 | 10460 |  |
| SLF II right | 0.755 | 14.08 | 1.18E-04 | -3.7472 | 0.2536 | 2.905 | -1.9566 | 3097.824 |  |
| SLF II left | 0.769 | 15.15 | 8.10E-05 | -4.5563 | -0.0293 | 3.3544 | -1.1772 | 3881.043 |  |
| SLF I left | 0.955 | 91.03 | 2.27E-09 | 8.6577 | -0.514 | -5.1696 | 4.2894 | -7612.7 |  |
| Tract | Adj R<sup>2</sup> | Fstat | Prob(Fstat) | β<sub>0</sub> | β<sub>1</sub>(ficvf) | β<sub>2</sub>(fiso) | β<sub>3</sub>(MD) |  |  |
| SLF I right | 0.898 | 50.92 | 8.89E-08 | -7.9241 | 6.0813 | -2.4927 | 6363.9379 |  |  |
| SLF II right | 0.538 | 7.589 | 2.98E-03 | -1.2137 | 1.1996 | -0.7505 | 992.3858 |  |  |
| SLF II left | 0.784 | 21.52 | 1.65E-05 | -4.8639 | 3.5599 | -1.3555 | 4141.768 |  |  |
| SLF I left | 0.942 | 92.83 | 1.77E-09 | 3.3861 | -1.6484 | 2.1134 | -3240.57 |  |  |
| Tract | Adj R<sup>2</sup> | Fstat | Prob(Fstat) | β<sub>0</sub> | β<sub>1</sub>(ficvf) | β<sub>2</sub>(MD) |  |  |  |
| SLF I right | 0.869 | 57.56 | 9.18E-08 | -3.6501 | 2.9309 | 2847.19 |  |  |  |
| SLF II right | 0.428 | 7.356 | 5.94E-03 | -0.3324 | 0.5072 | 273.7586 |  |  |  |
| SLF II left | 0.754 | 27.08 | 1.05E-05 | -2.2825 | 1.6053 | 2055.1872 |  |  |  |
| SLF I left | 0.897 | 74.8 | 1.58E-08 | 0.5043 | 0.2945 | -698.516 |  |  |  |

## Discussion

Our study presents a data analytical approach of integrating WBSI derived metabolite maps and NODDI/DTI derived reconstruction of SLF I and SLF II in healthy individuals. This method offers a novel, user-independent and comprehensive framework for evaluating spatially resolved metabolite and microstrusctural alterations along the distinct segments of SLF in healthy individuals. Mutual information data revealed WBSI derived white matter metabolite maps and NODDI/DTI derived parameters from distinct segments of SLF had strong spatial concordance and these two imaging modalities aligned well in their spatial distributions. Additionally, the tract profiles for metabolites and microstructure showed good repeatability as assessed from single subject data acquired three times. The inter-subject variability was also within acceptable ranges, as measures by the inter-subject CVs, with the free water fraction (fiso) exhibiting a relatively higher variability, a finding that was consistent with prior literature(Chung, Seunarine, & Clark, 2016). Our findings also demonstrate significant associations between WBSI-derived parameters and NODDI/DTI derived parameters along the course of the SLF. These results highlight the feasibility and utility of integrating WBSI and NODDI/DTI metrics for a comprehensive assessment of white matter pathways in SLF.

SLF plays a critical role in higher-order cognitive functions, including attention, language processing, and working memory. Using 1H-MRS and dMRI, the majority of prior studies (Catani et al., 2003; Cho et al., 2008; Michelutti et al., 2025; Su et al., 2016) have primarily focused on examining either the metabolic or microstructural integrity of SLF separately, neglecting the potential interplay and connection between these two characteristics.

While using a combined analytical approach involving DTI and single voxel 1H-MRS data in TBI, a study found significant diffusion and metabolite abnormalities after a concussion in adolescent athletes compared to control group. However, there were differences in the ROIs placement for 1H-MRS and DTI data analyses (Manning et al., 2017). In another study, significant differences in DTI derived parameters were observed between normal controls and schizophrenia patients from brain regions along the path of SLF. However, no significant differences in metabolite profiles were observed between the two groups (Rowland et al., 2009). While these studies have advanced our understanding of SLF functionality, they fail to capture the complex interplay between the structural integrity of white matter and metabolite alterations along the path of SLF. This gap in understanding is mainly attributed to the lack of overlap in the areas of interest studied by 1H-MRS and DTI.

The co-localized microstructure and metabolite tractometry based approach introduced here represents a significant methodological advancement. This allows for the extraction of region-specific tract profiles, enabling the evaluation of microstructural and metabolic parameters along the core of anatomically defined white matter tracts in tandem. Conventional ROI-based and voxel-wise approaches often suffer from losing the fine details of spatial information and anatomical correspondence, which could be well addressed by tractometry based methods. The use of tract-centric methodologies enhances the spatial precision of WBSI and addresses the confounding effects of partial volume contamination. An enhanced statistical power by averaging parameters along the centroid of a reconstructed tract is yet another advantage of tractometry based methods, which can reveal significant changes even when voxel-based analyses fail to detect such findings. This improvement likely arises from aggregating data across multiple noisy voxel-wise estimates rather than considering each voxel in isolation, thereby reducing variability and amplifying meaningful patterns (Jones & Nilsson, 2014). Such an approach would thus be helpful to detect a small, localized effect of a few voxels within a long-range white matter tract, especially occurring in the early stages of a neurodegenerative disorder.

The current study employed NODDI, an advanced dMRI technique for estimating the microstructural complexity of neurites (combination of dendrites and axons) and myelin integrity by employing multi-compartment models to describe various white matter features. Traditionally used DTI sequence models the diffusion process within each voxel as a single, ellipsoidal shape representing the dominant direction of water molecule movement. In contrast, NODDI models the diffusion in each voxel as a combination of three distinct compartments and these are: intracellular (restricted anisotropic non-Gaussian diffusion), extracellular (hindered anisotropic Gaussian diffusion), and CSF (isotropic Gaussian diffusion). NODDI-derived parameters include intra-cellular volume fraction (ficvf, reflects packing density of axons or dendrites); orientation dispersion index (ODI, a measure of orientational coherence of neurites); and isotropic volume fraction (fiso, estimates the extent of CSF space) respectively (Caverzasi et al., 2016; Zhang, Schneider, Wheeler-Kingshott, & Alexander, 2012). Some studies have demonstrated that NODDI provides a more detailed and accurate assessment of white matter integrity compared to DTI. Instead of using single or multivoxel conventional spectroscopic sequences that are constrained by limited brain coverage, the WBSI sequence that provides high-resolution metabolic maps (voxel size =0.10 cm^3^) covering both supratentorial and infratentorial brain regions was used in the present study (Ebel, Soher, & Maudsley, 2001; Maudsley et al., 2006) These maps can be spatially co-registered to anatomical images facilitating mapping of metabolite alterations from multiple brain regions with a less probability of partial volume averaging. Typically, a large number of voxels (∼ 16000) are obtained, potentially obviating the subjectivity and user bias for placing voxels in a tissue of interest.

Metabolic and microstructural characteristics of tissues are closely related, and understanding their correlations can provide valuable insights into disease mechanisms and progression. Choline (Cho) is not just a structural component of myelin but also actively participates in the cellular processes that drive myelin formation, maintenance, and repair. The Cho levels (Cho/Cr and Cho/NAA) are uniformly distributed in the white matter regions (Pouwels & Frahm, 1998), and their concentrations are known to be greater in white matter than in grey matter (Li et al., 2022; Nitsch et al., 1993), likely reflecting normal membrane turnover and cellular density in these regions (Rae, 2014). While ficvf reflects the proportion of intracellular space within a voxel and higher ficvf indicates greater neurite density, FA signifies the degree of diffusion anisotropy present within a voxel and higher FA values indicates well-organized, and myelinated white matter fibers in the brain. Several factors such as cellular packing, the presence of intracellular organelles, cell membranes, and macromolecules determine the MD values. Moreover, alterations and redistributions of water molecules between intracellular and extracellular tissue compartments are also known to influence MD values. Several earlier studies have demonstrated significant associations between 1H-MRS and dMRI derived parameters in multiple neurological disorders(Dennis et al., 2018; A. A. Maudsley et al., 2015). To accurately interpret such deviations along the path of SLF, it is essential to first establish normative patterns of association between parameters derived from these two co-localized imaging modalities in healthy individuals within the same brain regions. This baseline understanding of normal variation will facilitate identification of atypical patterns present in neurological conditions in which SLF is known to be compromised.

While investigating the metabolic and microstructural integrity of SLF, a significant positive association between FA, ficvf and Cho/Cr ratio with a concomitant negative relationship between MD and Cho/Cr was observed in the present study. This suggests that a higher membrane metabolism and cellular density is directly linked to greater microstructural coherence and white matter integrity. Conversely, an inverse relationship with MD in the same regions is consistent with the notion that an increased membrane turnover corresponds to more restricted diffusion, indicative of denser tissue architecture. Similar, but less prominent associations between Cho/NAA ratio and FA/MD were observed in the present study. These findings suggest that cellular metabolic activity is closely associated with structural organization of white-matter fibers in SLF.

Despite presenting a novel analytical approach, our study is limited by some shortcomings. Keeping a simplified approach in mind, only two metabolite ratios (Cho/Cr and Cho/NAA) were considered in the present study for an initial proof-of-concept investigation. This limited scope allowed focused analysis and easier interpretation of the initial findings. Other brain metabolites such as myo-inositol (mI) and Glutamate + Glutamine (Glx) were not examined in this study. Although WBSI covered the entire brain, only 60-70% of whole-brain voxels were of good quality. In this study, poor-quality spectra occurred mainly in the inferior temporal lobe and brainstem regions. To eliminate bias from this unreliable data, our spatial analysis was restricted to SLF I and II tracts, excluding SLF III and IV.

In conclusion, our proposed methodological advancement of combining WBSI and NODDI/DTI derived parameteric maps within an integrated analytical framework offers a more comprehensive, reliable and user-independent approach to understanding regional metabolite patterns and white matter integrity along the path of SLF. In future, this novel framework can be utilized for longitudinal cohorts and clinical trials evaluating therapeutic efficacy. Moreover, this method can be used for establishing varying levels of associations between metabolite and microstructural alterations from different segments of SLF and clinical measures in neurological disorders affecting SLF integrity.

## Acknowledgements

We thank all the normal, healthy individuals for their participation in this study as volunteers.

## Data Availability Statement

The data that support the findings of this study are available from the corresponding authors [S.C. and S.M.] upon reasonable request.

## Ethics Statement

All procedures performed in this study involving human participants were in accordance with the ethical standards of the institutional research committee and with the 1964 Helsinki Declaration and its later amendments or comparable ethical standards. This study was approved by the Institutional Review Board (IRB# 817383) and was compliant with the HIPAA (the Health Insurance Portability and Accountability Act).

## Consent

Written informed consent was obtained from each participant prior to enrollment in this study.

---

**Figure S1.**
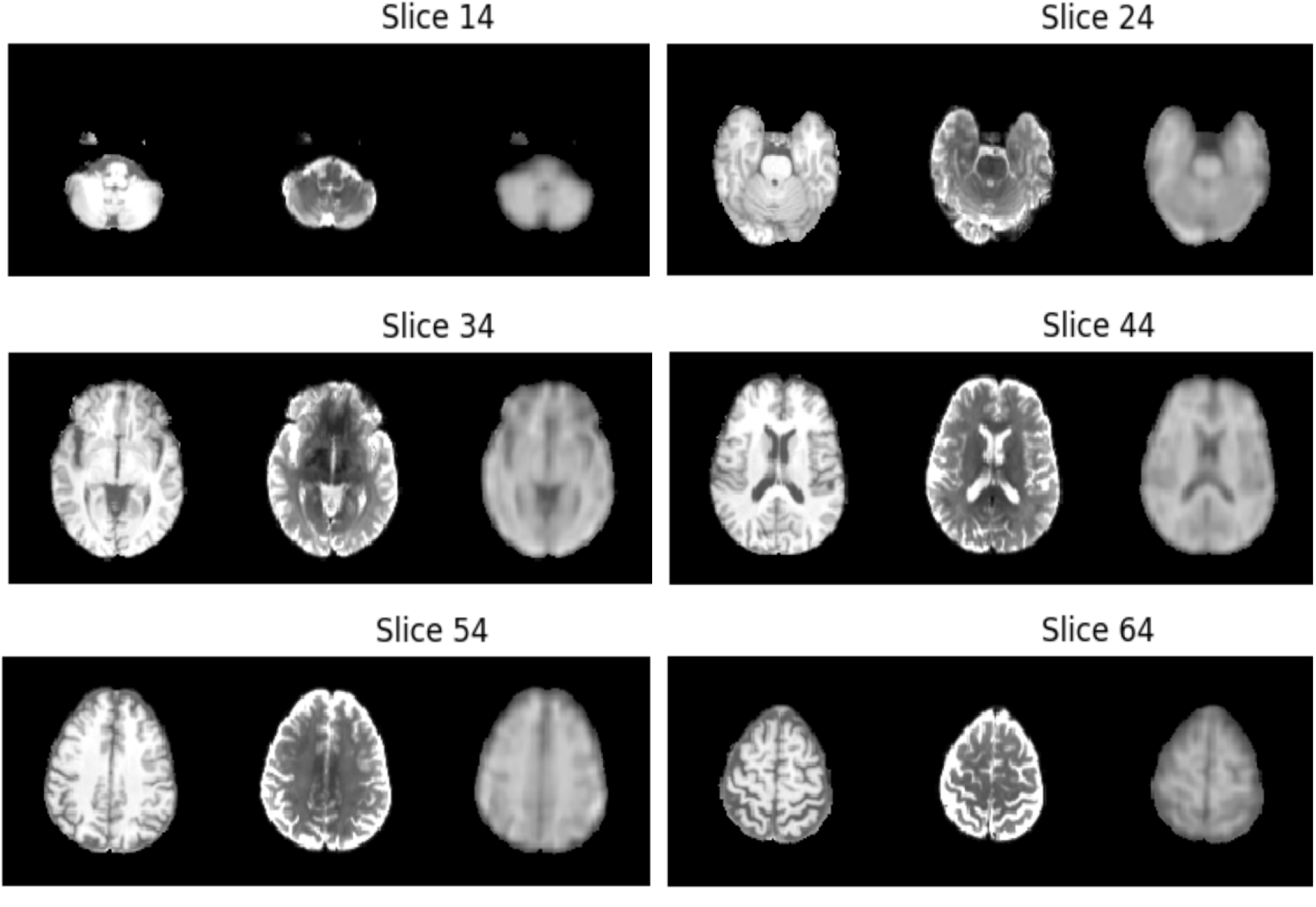
Co-localization across modalities in a representative subject. From left to right are shown the coregistered T1, the b0, and the coregistered WaterSI images, for a few exemplary axial slices.

**Figure S2.**
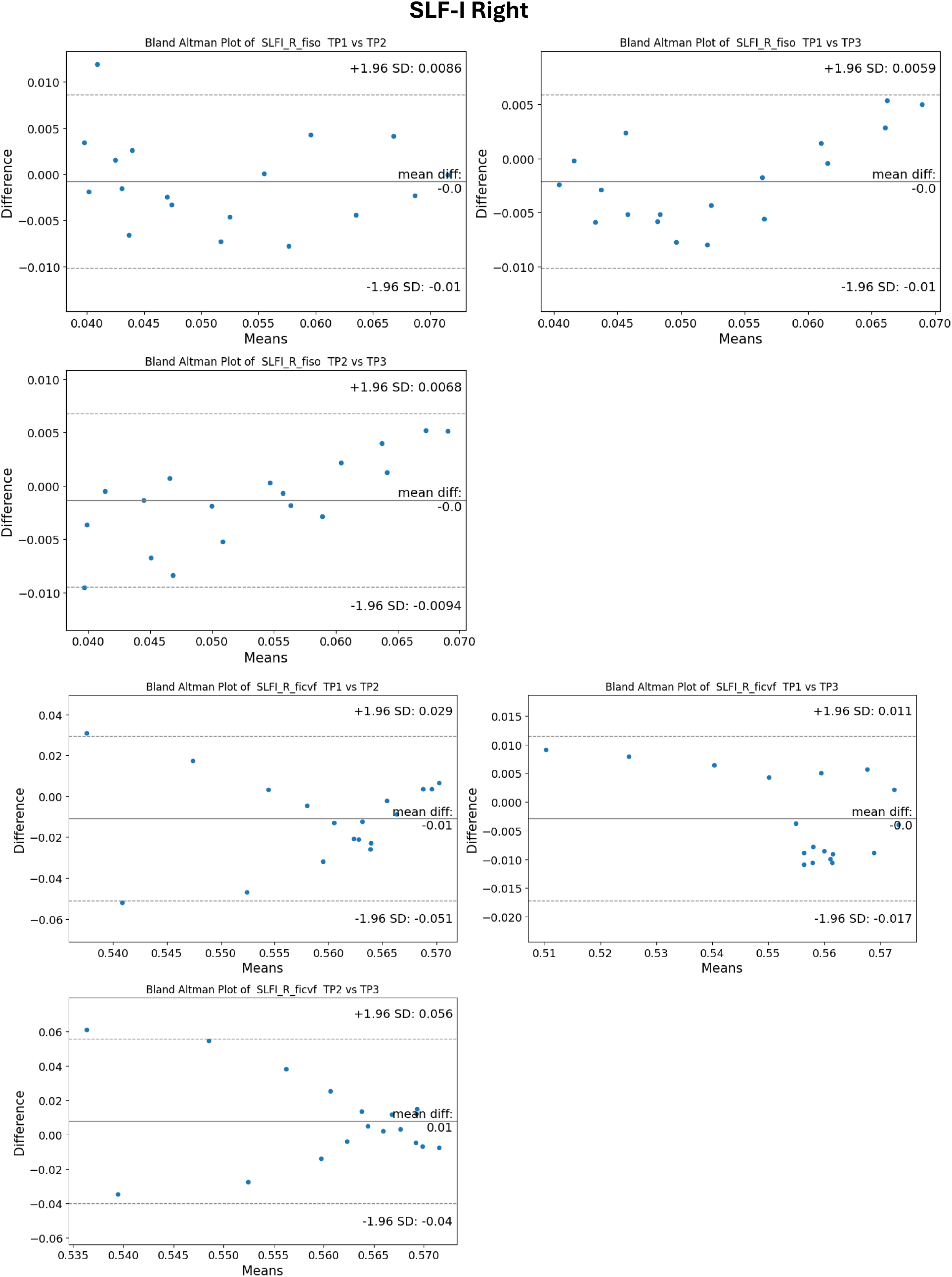
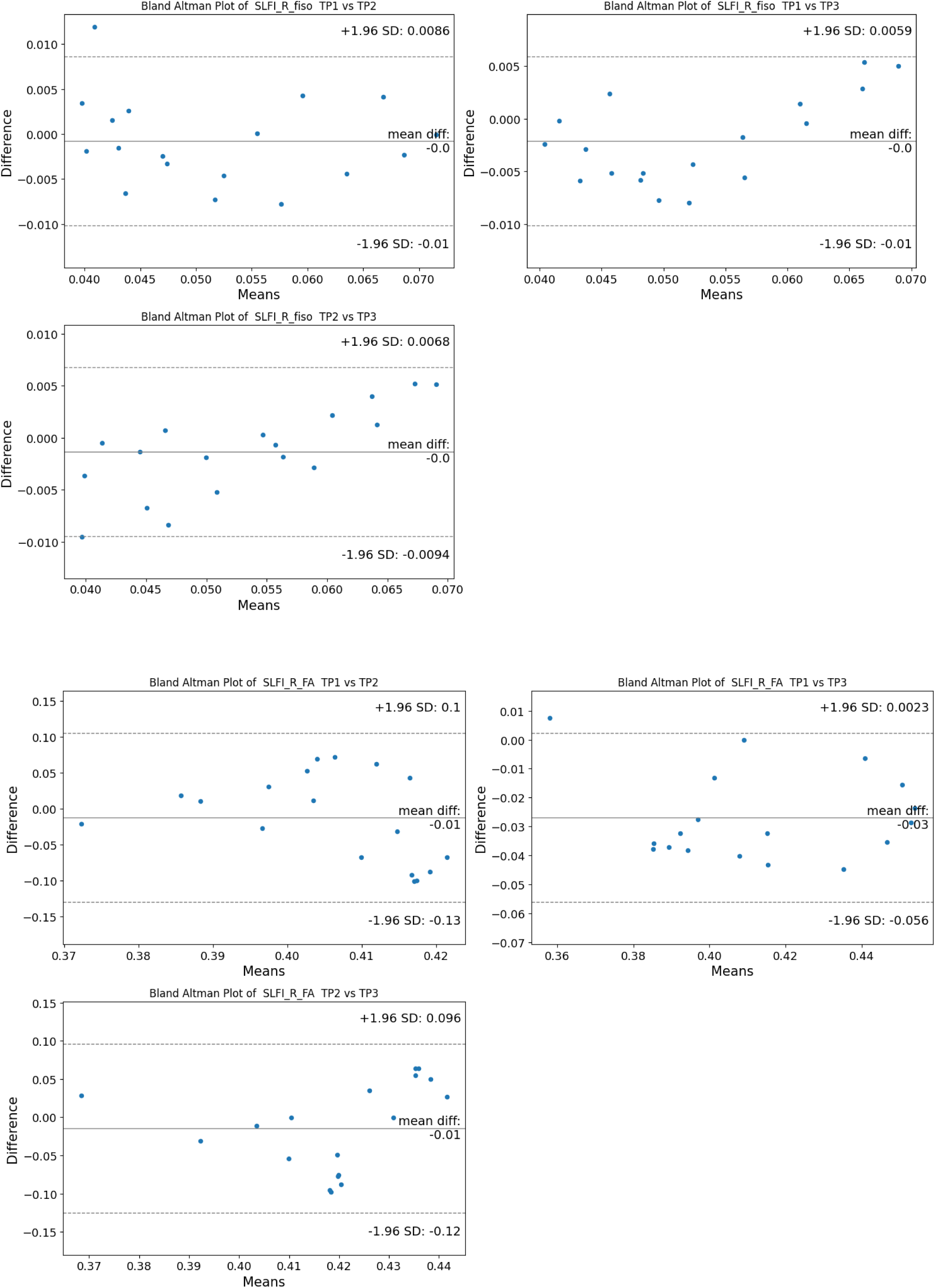
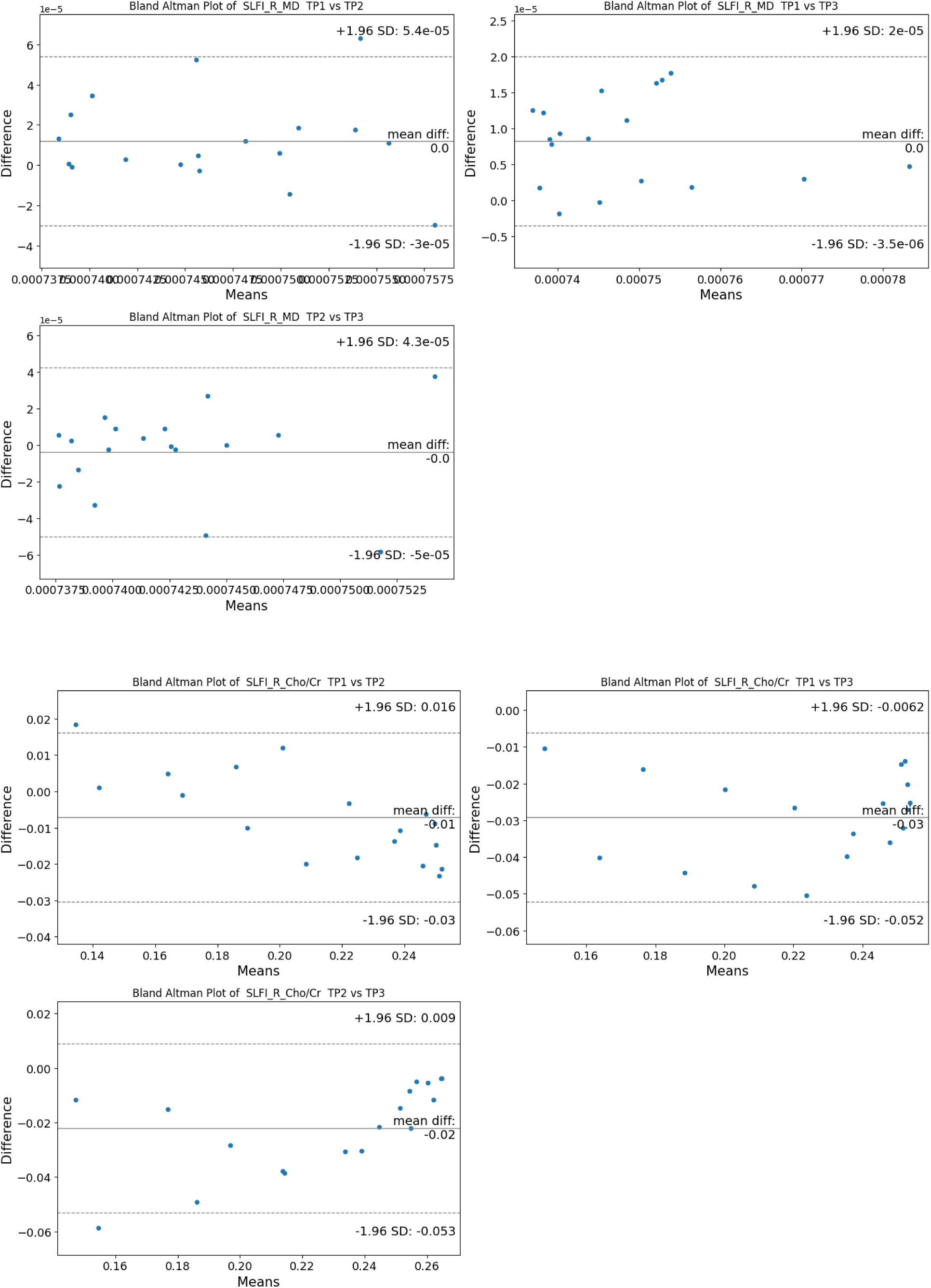
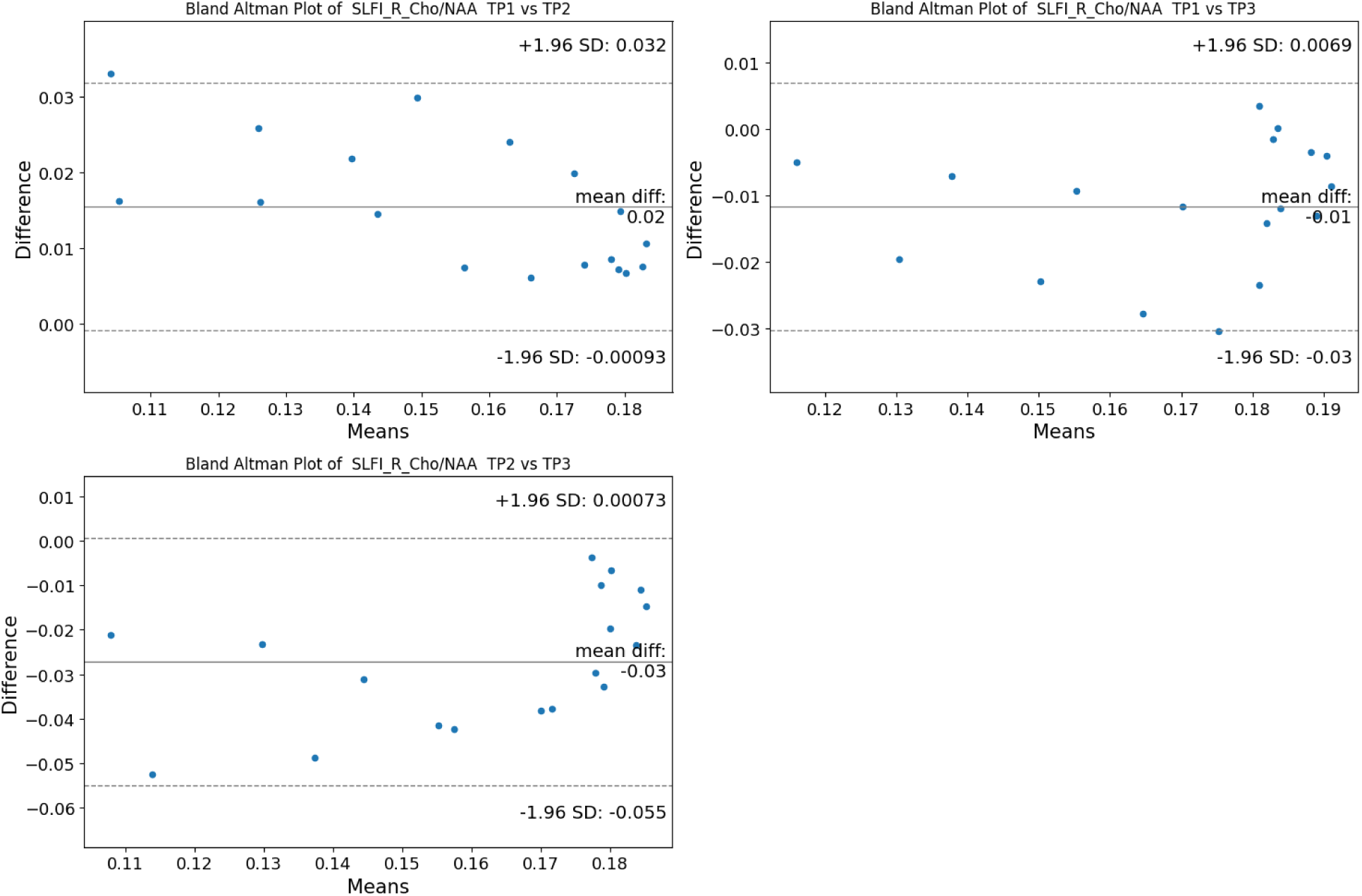
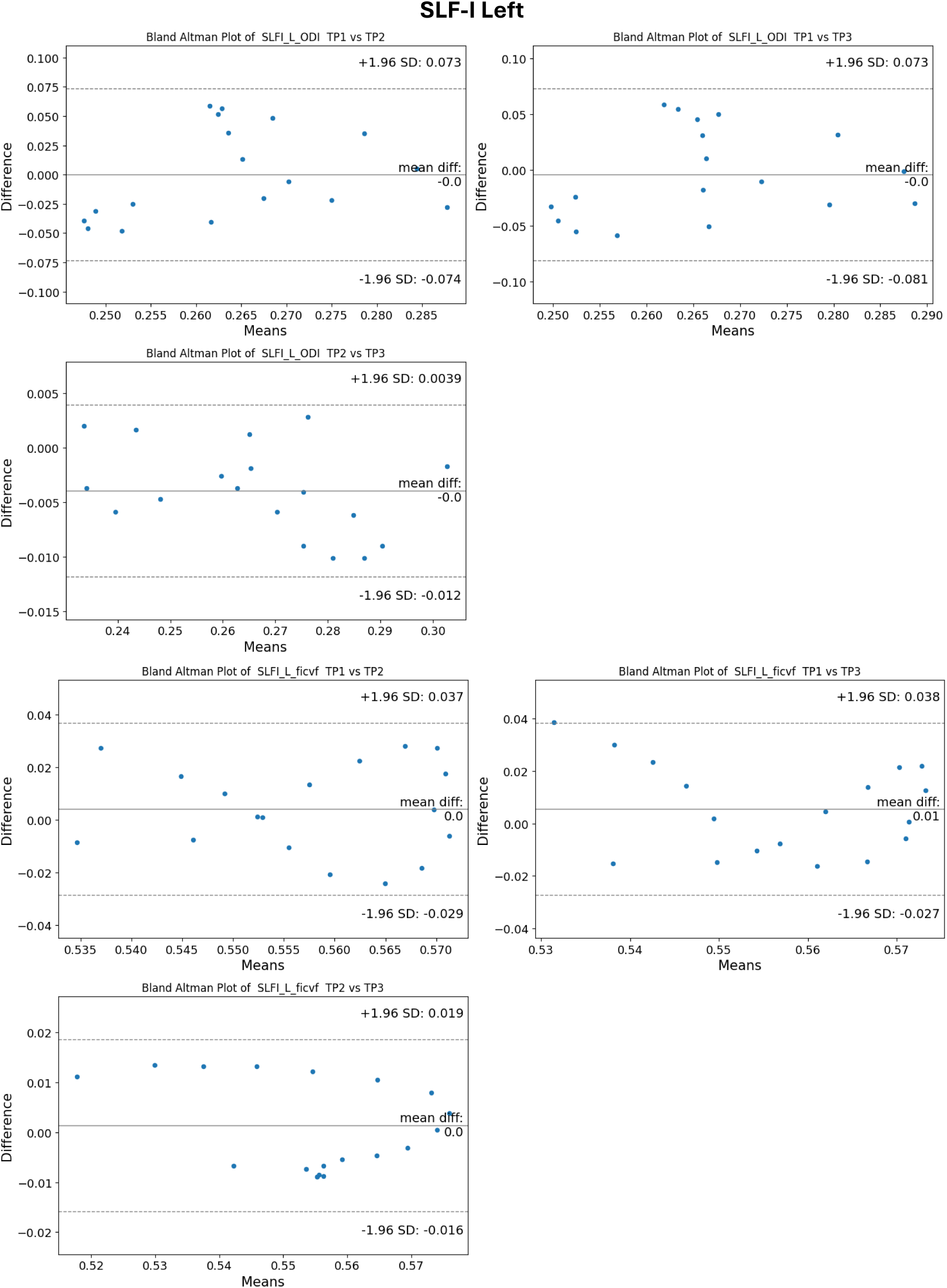
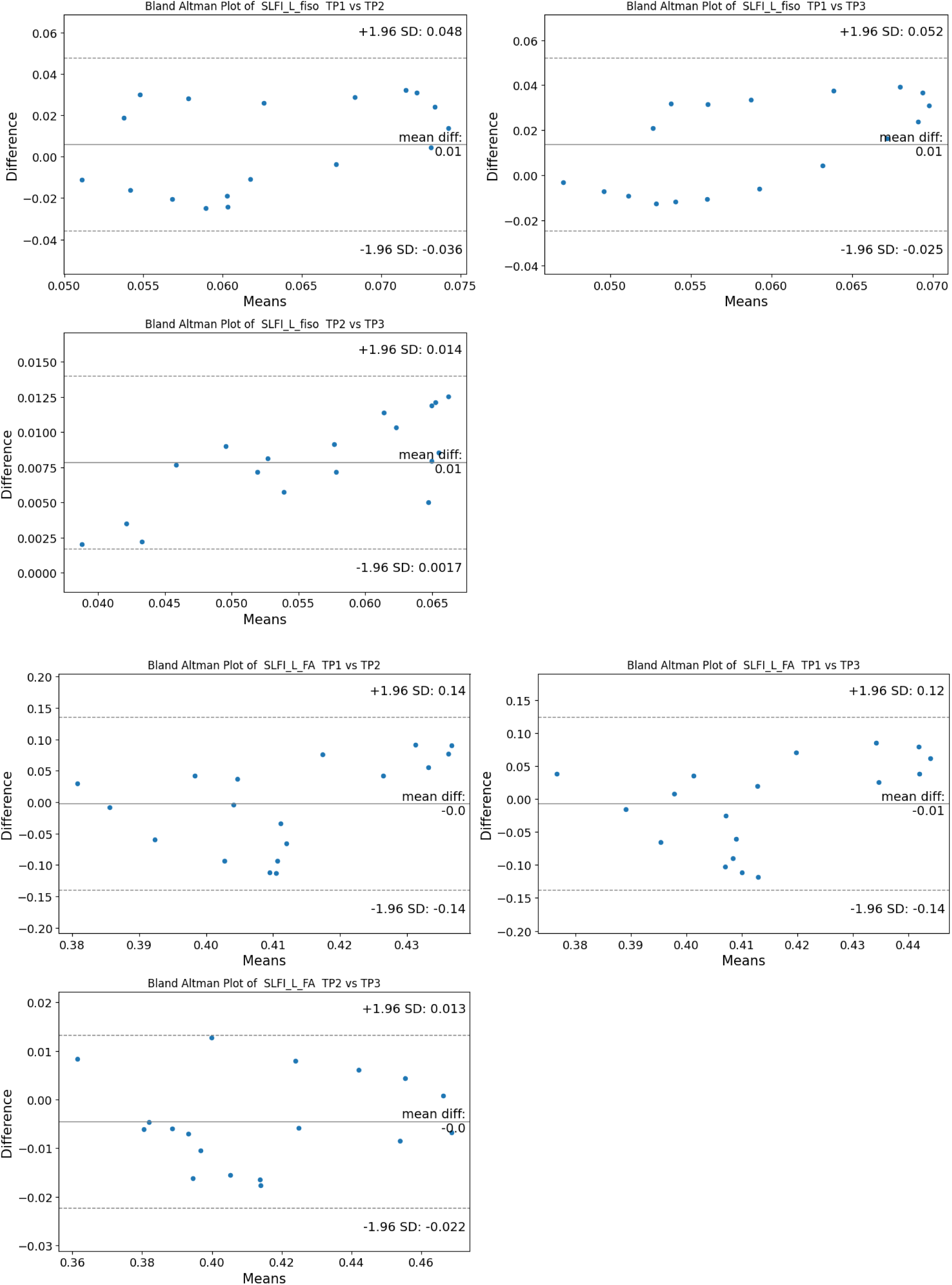
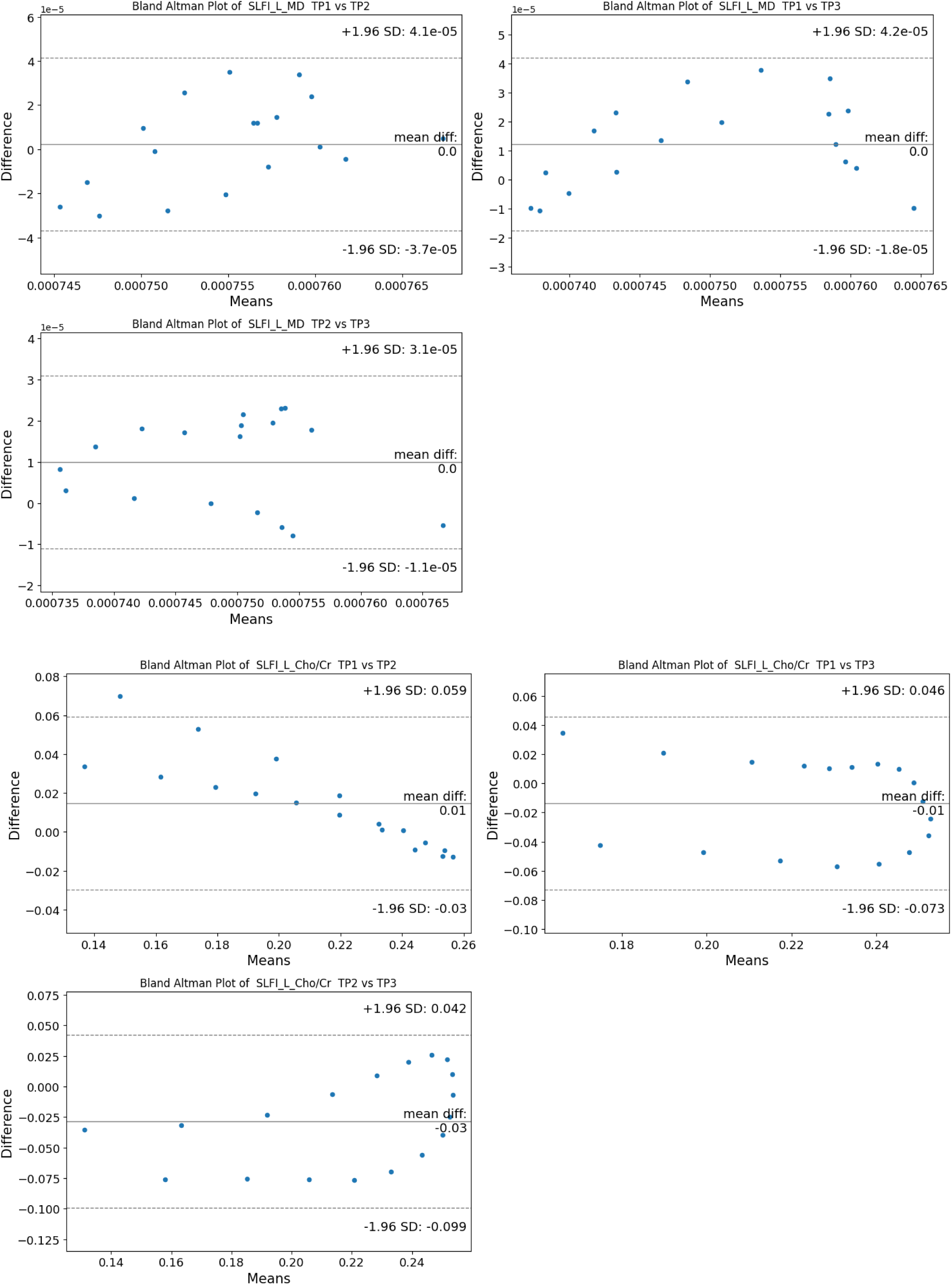
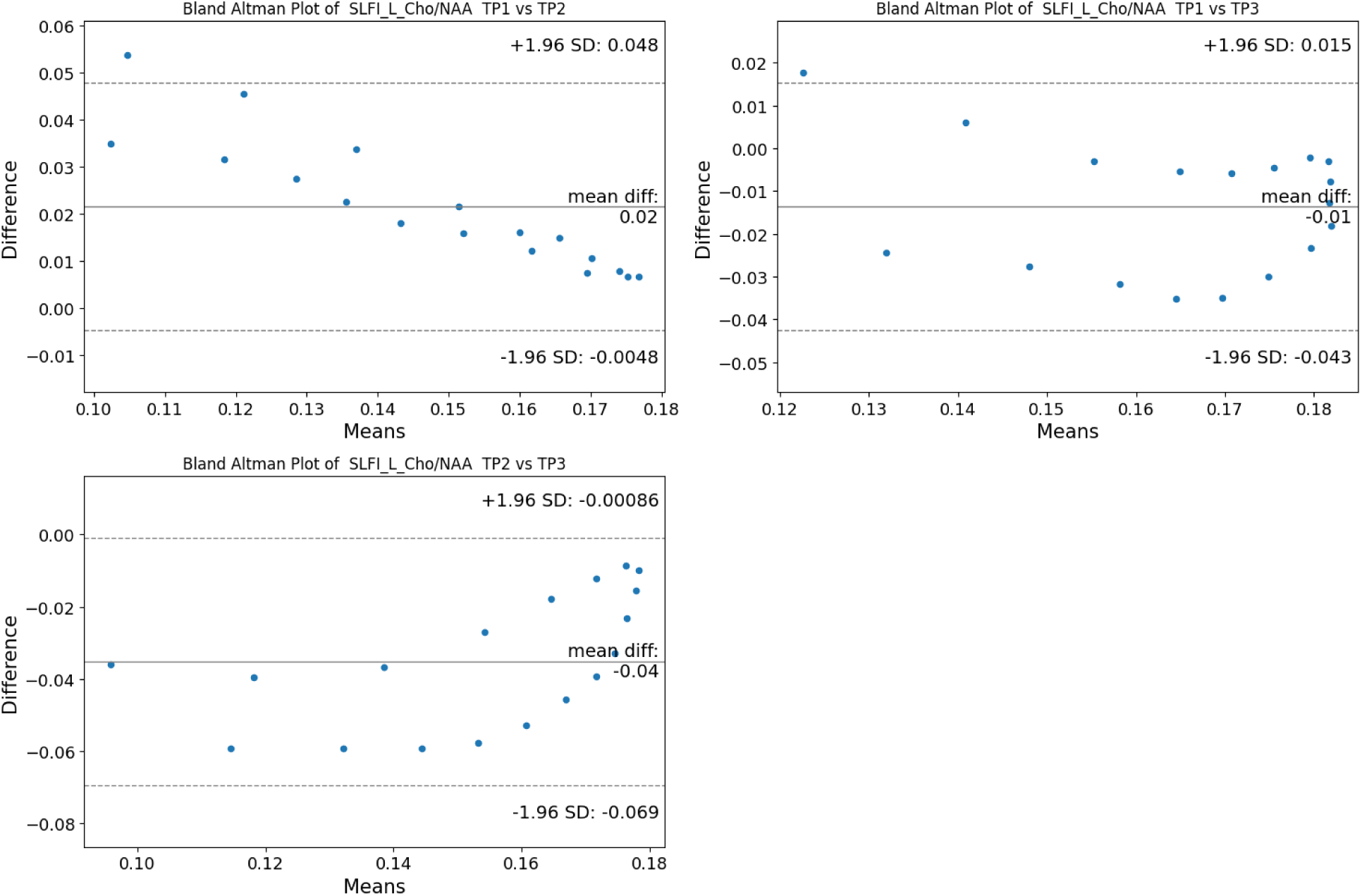
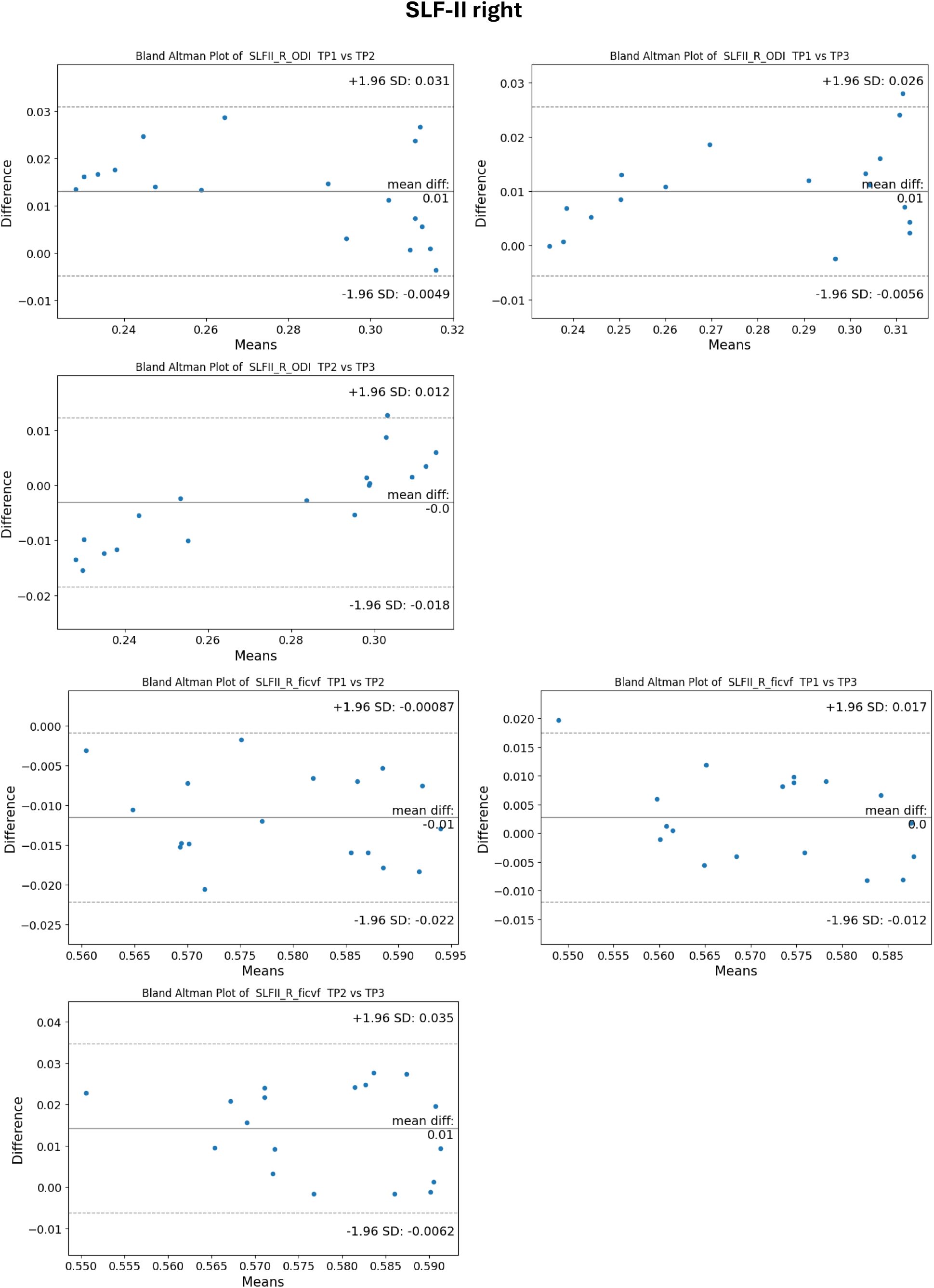
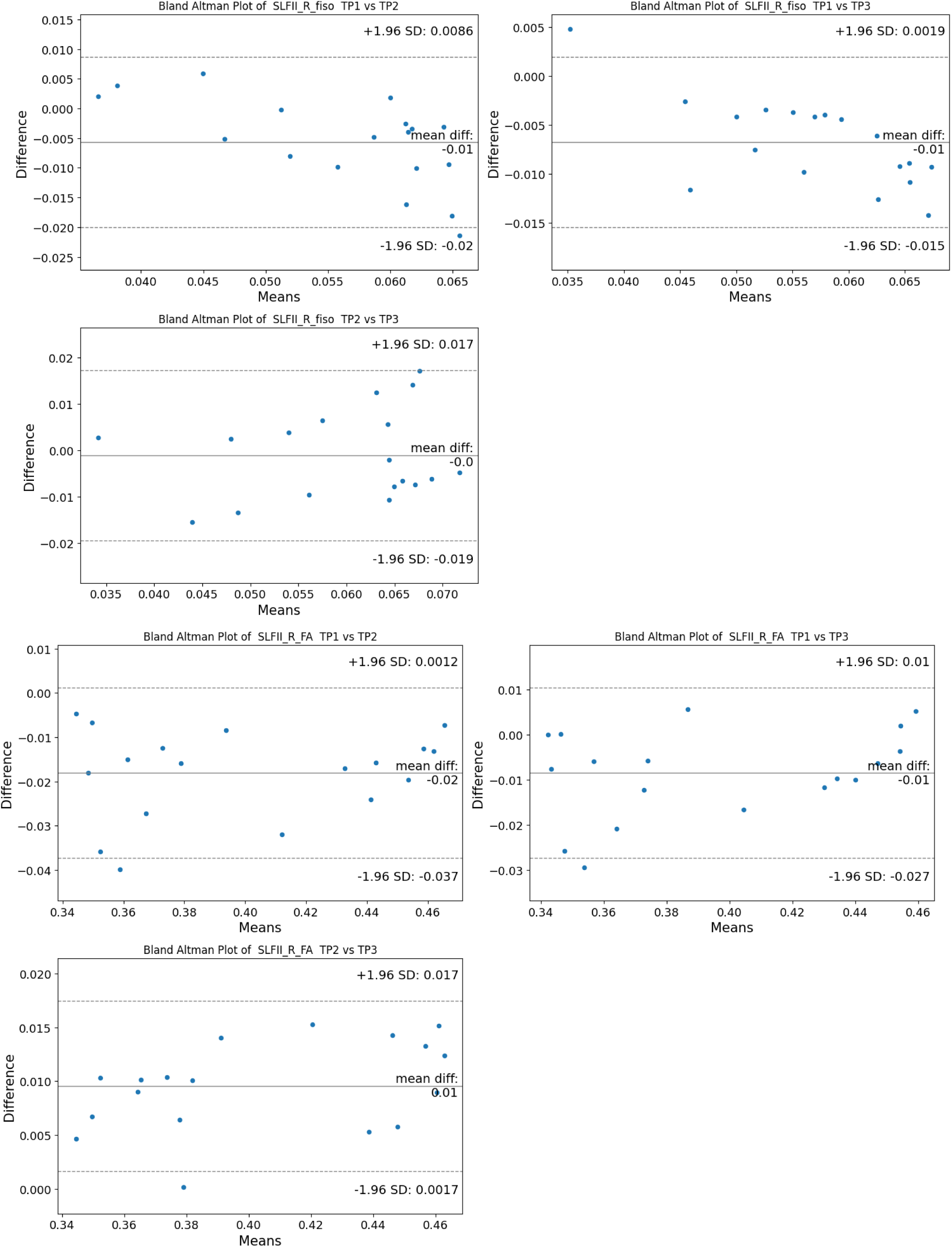
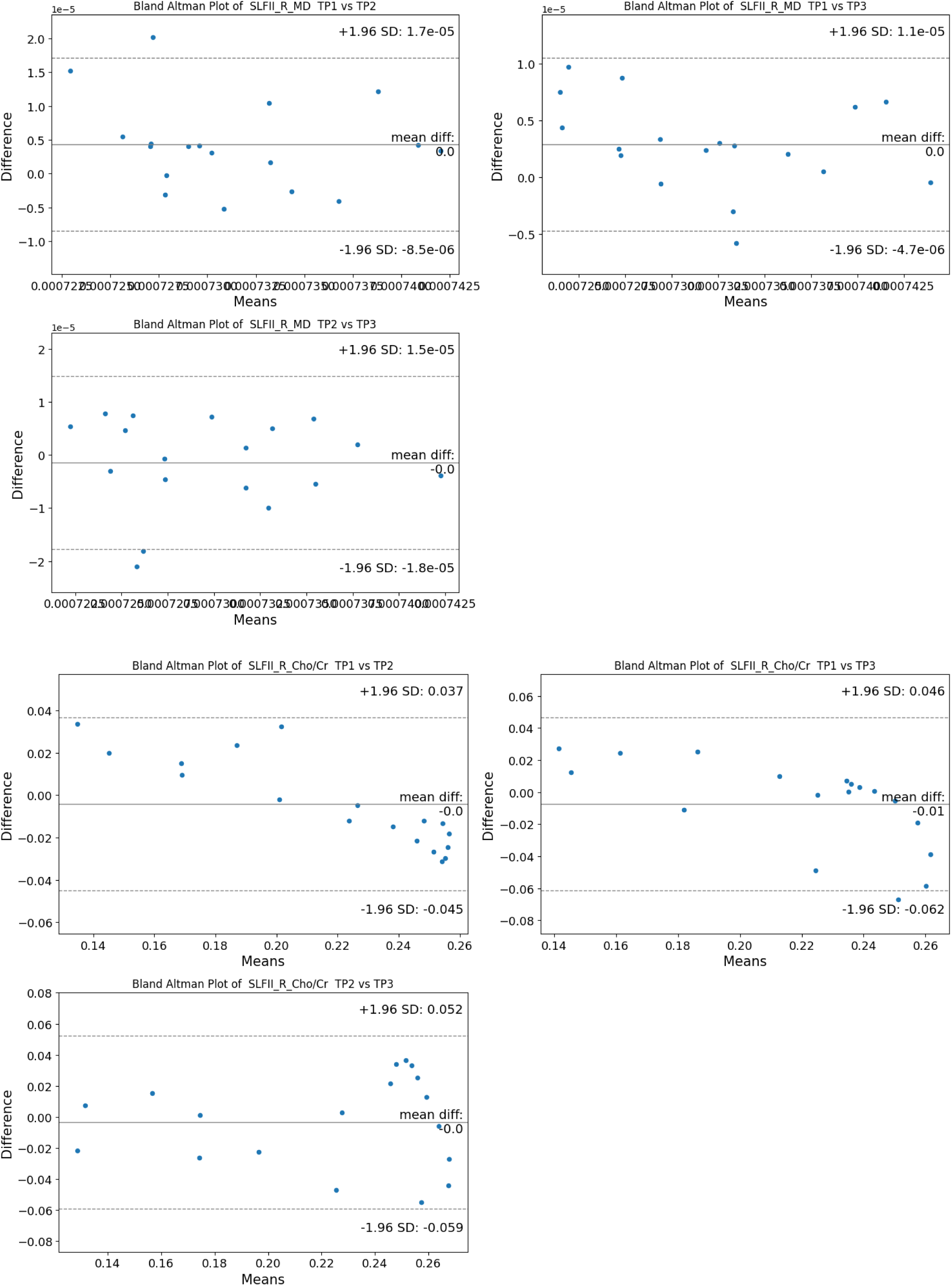
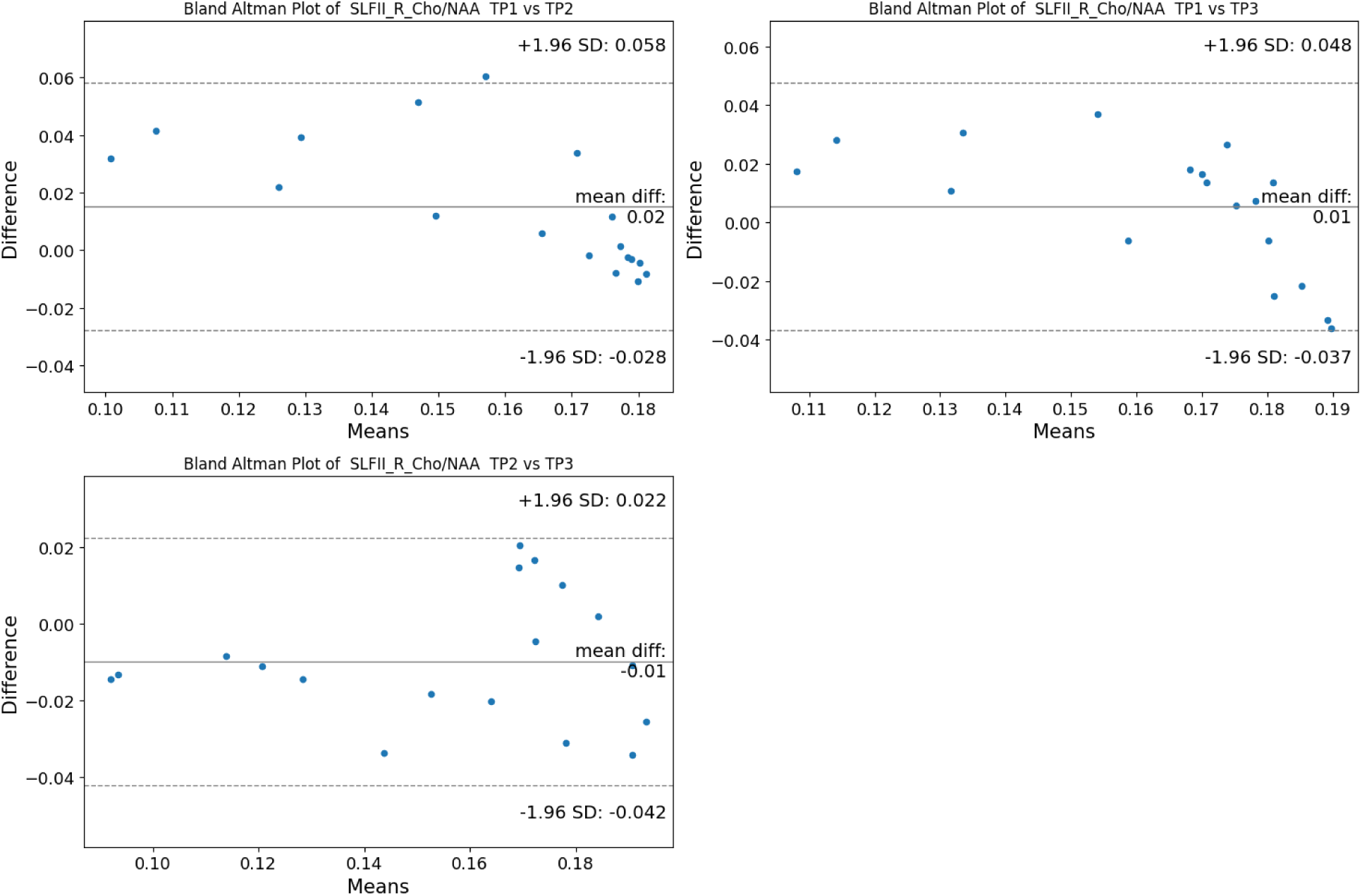
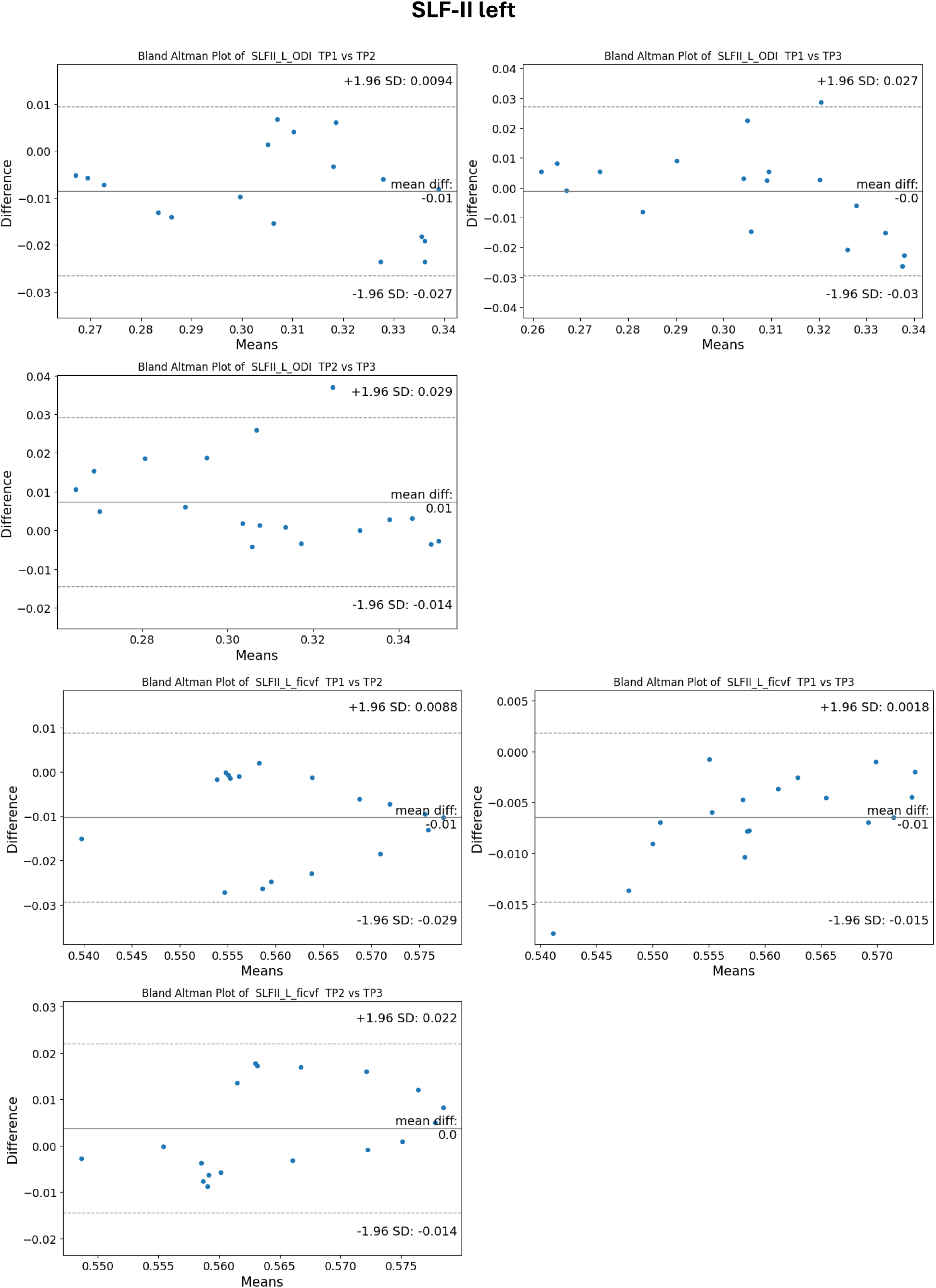
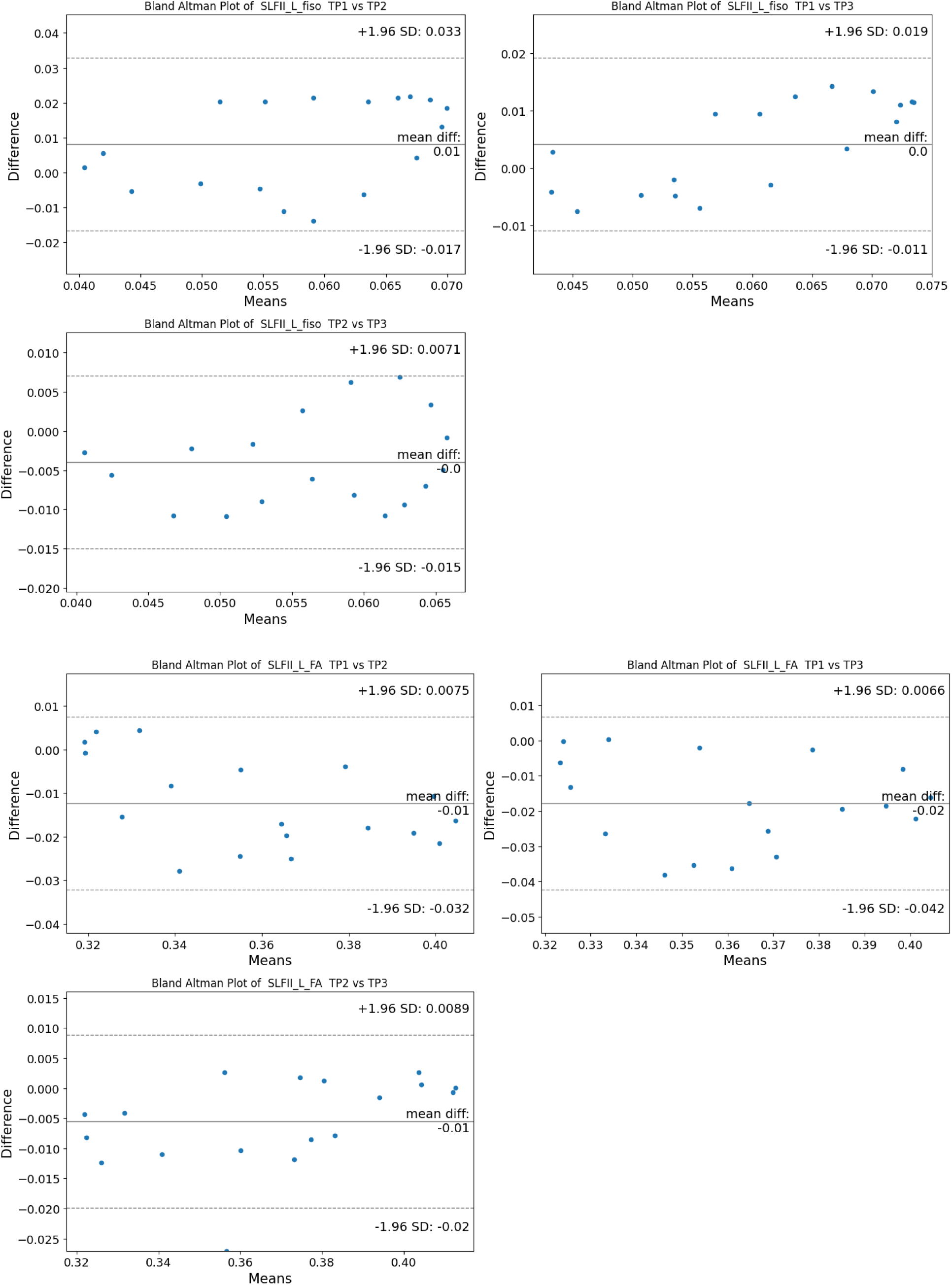
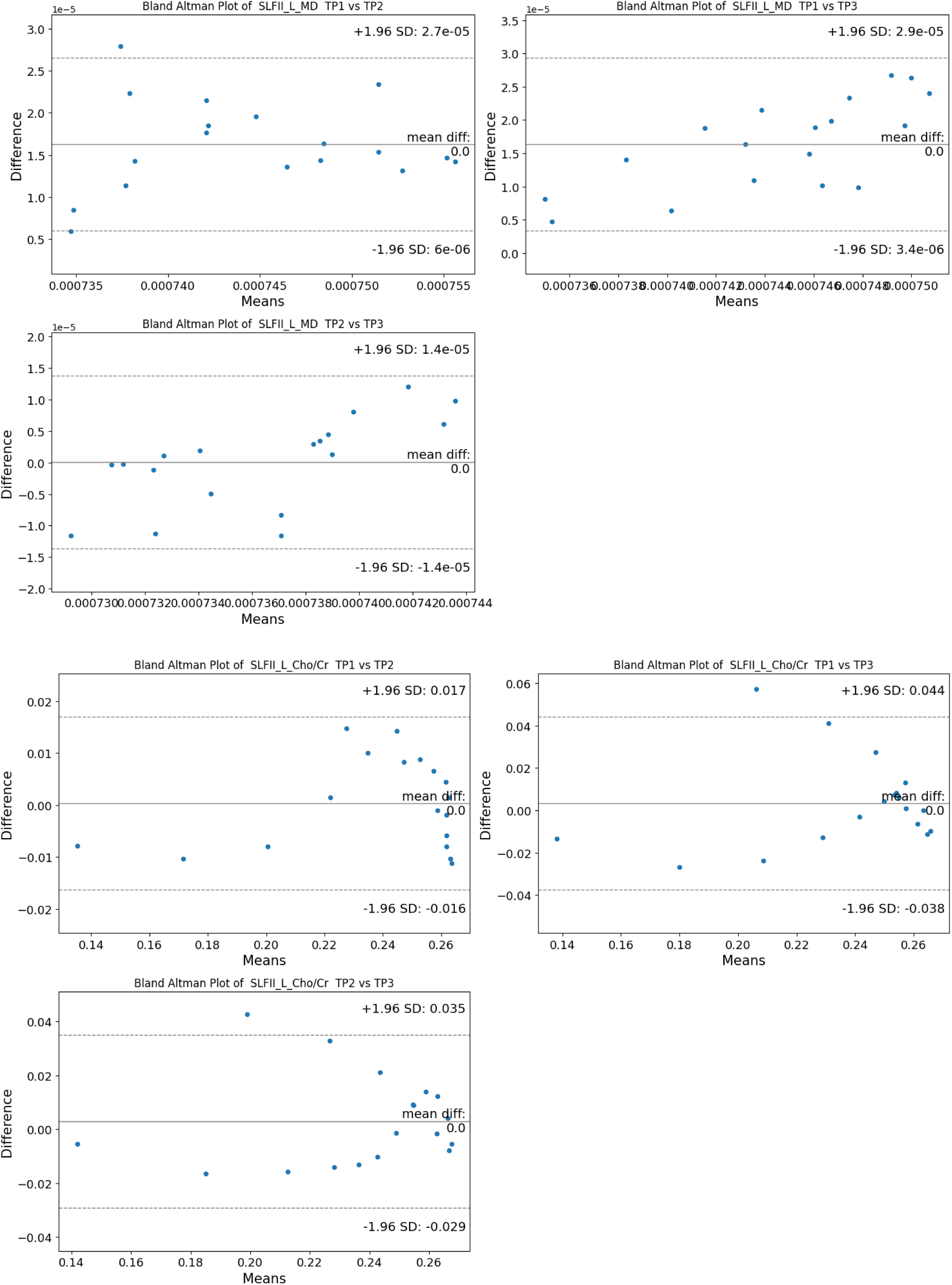
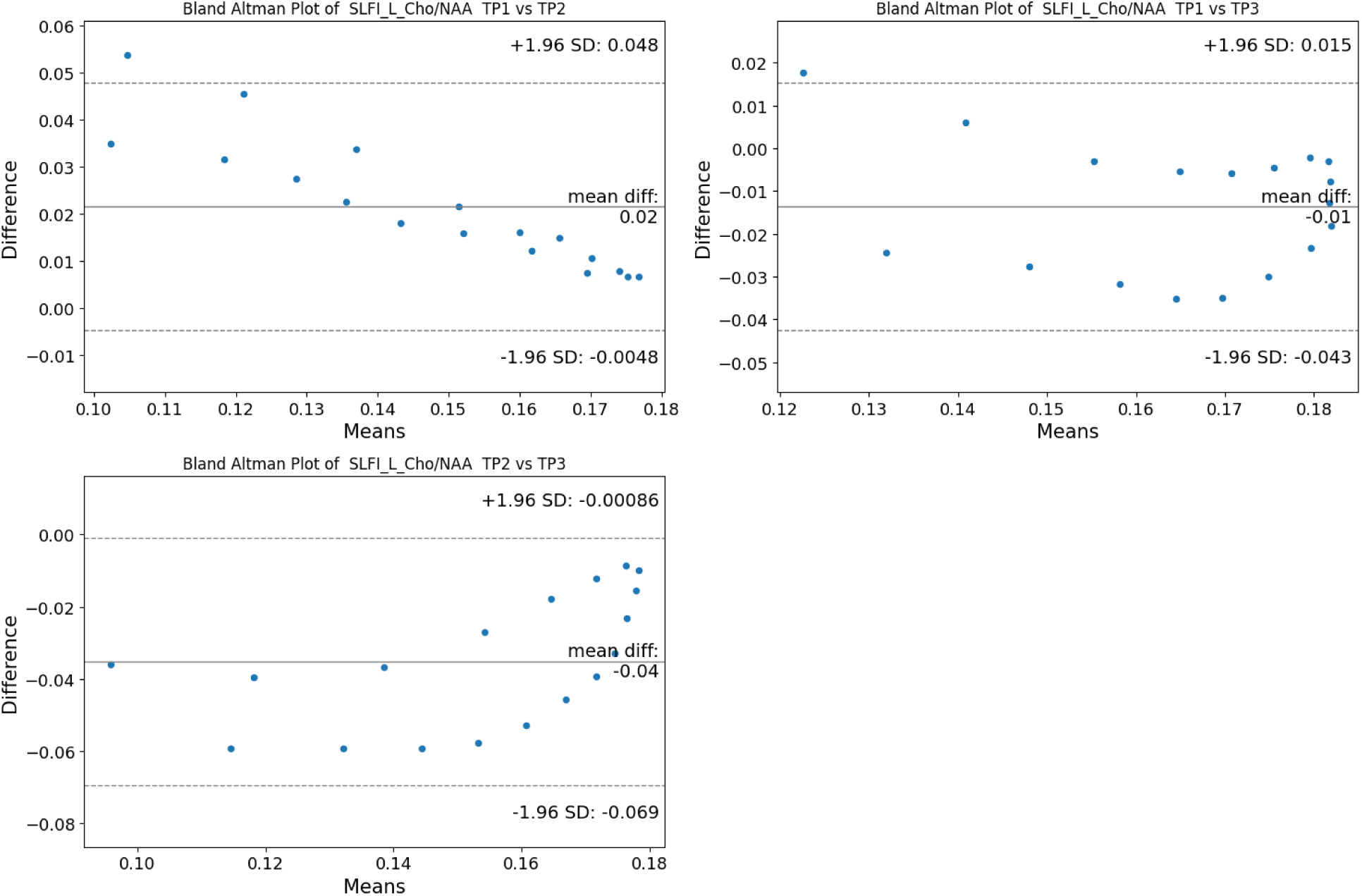
Bland Altman Plots for the different SLF segments

